# Blood-based RNA-Seq of 5412 individuals with rare disease identifies new candidate diagnoses in the National Genomic Research Library

**DOI:** 10.64898/2026.03.19.26348811

**Authors:** Jenny Lord, Alistair T Pagnamenta, Letizia Vestito, Susan Walker, Carolina Jaramillo Oquendo, Anthony EF McGuigan, Alexander Ho, Christopher Odhams, Julius OB Jacobsen, Sarju Mehta, Evan Reid, Mary O’Driscoll, Christopher M Watson, Laura A Crinnion, Rachel L Robinson, Hannah Musgrave, Richard J Martin, Terena P James, Mark T Ross, Marianna Kyritsi, Leonardo Carnielli, Nicholas Walker, Dunja Vucenovic, Uma Maheswari, Francisco E Baralle, Jenny C Taylor, Jamie M Ellingford, Dalia Kasperaviciute, Lily Hoa, Greg Elgar, Matthew A Brown, Damian Smedley, Diana Baralle

## Abstract

RNA sequencing (RNA-Seq) is increasingly used alongside exome and genome sequencing to identify causal variants underlying rare Mendelian disorders. We present short-read RNA-Seq data from 5,412 individuals with a diverse range of rare disorders recruited to Genomics England’s 100,000 Genomes Project. We show that the proportion of genes from gene panels applied to different disorders which are well captured (transcripts per million (TPM) ≥ 5) from blood RNA varies widely, highlighting differences in applicability across disorder types. Using OUTRIDER and FRASER2 to identify gene expression and splicing outliers respectively, we identify at least one outlier event in a disorder relevant gene in 20% of the cohort. To prioritise likely diagnostic candidates, we apply multiple strategies including focussing on outlier events in known haploinsufficient genes (n=78), integrating outliers with structural variant calls (n=19), and using strategies integrating phenotypic presentation (Exomiser, n=39). We present a series of candidate diagnoses involving diverse variant types and disease mechanisms, demonstrating the broad utility of RNA-Seq in identifying and prioritising diagnostic candidates in individuals with a variety of different rare conditions and no known genetic diagnosis. Our findings demonstrate that blood-based RNA-Seq can deliver clinically relevant findings across a broad range of rare disorders.

## Introduction

Rare disorders each individually affect fewer than 1 in 2,000 individuals, but with over 8,000 different disorders thought to exist, their cumulative frequency is around 1 in 17 (GeneticAllianceUK). It is thought that most rare disorders are genetic, and advances in exome and genome sequencing (GS) have greatly improved diagnosis. However, even with GS, more than half of patients remain without a genetic diagnosis (Edsjo et al, 2023; Investigators et al, 2021). Many factors are likely to contribute to this, including incomplete knowledge of disease-linked genes, technical difficulties sequencing particular variant types, more complex modes of inheritance such as polygenic inheritance, environmental involvement in disease pathogenesis, and challenges in interpreting variants that disrupt splicing or occur in non-coding genomic regions.

Current diagnostic pipelines focus heavily on protein coding variants, with under-representation of splicing (Lord et al, 2019) and non-coding variants (Ellingford et al, 2022) in databases such as ClinVar (Landrum et al, 2018). There have been major improvements in the accuracy and throughput of splicing prediction tools in recent years, including SpliceAI (Jaganathan et al, 2019) and Pangolin (Zeng & Li, 2022), along with the development of guidelines for the interpretation of splicing (Sullivan et al, 2025) and non-coding variants in the clinical setting (Ellingford et al, 2022). Nonetheless, prediction approaches for splicing and non-coding variants remain imperfect, often necessitating costly functional validation (Brnich et al, 2019), and many pathogenic variants continue to be overlooked or reported as variants of uncertain significance.

RNA sequencing (RNA-Seq) offers a powerful complementary approach by directly profiling the transcriptome, enabling the detection of splicing disruptions and altered transcript abundance (e.g. due to copy number variants (CNVs) or regulatory variants) to be directly observed and quantified. The approach has been successfully applied in a variety of clinical contexts, with improvements in diagnostic yields ranging from 7.5 to 36% (Cummings et al, 2017; Dekker et al, 2023; Frésard et al, 2019; Gonorazky et al, 2019; Kremer et al, 2017; Oquendo et al, 2024; Yepez et al, 2022). However, these studies have typically been small and often focussed on specific phenotypes. Due to tissue specific differences in gene expression and splicing, a variety of source tissues have been explored, including RNA derived from muscle biopsy and fibroblasts derived from skin biopsies, often informed by the disorder of interest. The invasiveness, time and cost of obtaining such tissues limit scalability and clinical adoption. Consequently, there is strong rationale for exploration of the use of blood-based RNA-Seq across a broad range of disorders.

Here, we present findings from analyses involving whole-blood based total RNA-Seq data generated for over 5,000 individuals with rare disorders recruited to Genomics England’s 100,000 Genomes Project (100kGP). We performed systematic identification of gene expression and splicing outliers, identifying significant outlier events in potentially disease relevant genes in 20% of participants. We prioritise and explore likely diagnostic candidates in genes relevant to each individual’s phenotype and use Exomiser and overlap with structural variants to identify additional candidate diagnoses. Our results demonstrate that blood-based RNA-Seq is a scalable and clinically feasible approach that captures diverse pathogenic mechanisms across a wide range of rare disorders.

## Methods

### Cohort, sample collection, preparation and sequencing

At the time of recruitment to the 100kGP, many participants provided additional PAXgene whole blood samples for RNA extraction, in addition to samples for DNA extraction for GS. 5,701 participants were selected for RNA sequencing, the majority of whom were undiagnosed at the time of selection. Total RNA was isolated using the PAXgene Blood miRNA Kit (Qiagen, Germany) on the QIAcube Connect automated Nucleic Acid Extractor (Qiagen, USA) according to manufacturer’s protocol at UK Biocentre (Milton Keynes). The libraries were prepared from 100 ng total RNA by Illumina (Granta Park, England) with the Illumina Stranded Total RNA Prep, Ligation with Ribo-Zero Plus kit, which depletes globin and ribosomal RNAs plus additional custom deletion probes. Sequencing was conducted by Illumina using NovaSeq 6000 sequencers, generating 100bp paired-end reads. Sequence data were generated for 5,622/5,701 samples. 5,546 of these were successfully paired with a participant’s GS data after stringent integrity checks, and of these, 5,412 passed all quality and depth thresholds to form the final data set. This represents an overall pass rate of >95% for RNA samples submitted for sequencing.

Information on the cohort demographics including declared phenotypic sex, inferred karyotypic sex, genetically inferred ancestry, age at recruitment (and thus age at sampling), high level disorder classification and (Human Phenotype Ontology) HPO terms were obtained from LabKey (Web Resources). Gene panels from PanelApp (Martin et al, 2019) had been assigned to participants based on HPO terms reported as present as part of the 100kGP pipeline, these gene panel assignments were also obtained from LabKey, with up to date panel versions applied at the time of initial analysis (November 2023).

All 100kGP data (genome sequence and RNA-Seq, phenotype information, etc.) are stored in the National Genomic Research Library (NGRL, Web resources (2024)) and are accessible to registered researchers. All data presented in this paper have been exported from the NGRL following the guidelines set out by Genomics England for data export via the AirLock. To protect against re-identification, where HPO terms are given for specific participants, these have been abstracted (moving up the HPO hierarchy) such that the combination of terms given cannot be used to identify a group of fewer than five participants. As such, it is possible no participant with the specific combination of HPO terms given in this paper exists in the NGRL.

### Alignment, quantification and QC

Alignment of the RNA-Seq data to GRCh38, gene and transcript level quantification (GENCODE v32) and detection of fusion transcripts were performed using Illumina’s DRAGEN pipeline (v3.8.4), with implementation of STAR aligner (Dobin & Gingeras, 2015) and Salmon gene/transcript quantification (Patro et al, 2017).

A “light-touch” data QC approach was taken to ensure as many individuals as possible with potentially informative RNA-Seq would be included in the analysis set and thus have the potential to receive a diagnosis from the data.

RNASeQC2 (Graubert et al, 2021) was used to generate quality metrics on the data, and samples were included in the final analysis set if they passed the following conditions:

● Total reads >= 120M
● Percent correct strand reads > 97%
● Percent unmapped reads < 5%
● Average inner distance >= -40 bp
● Percent spliced reads >= 5%
● Percent rRNA reads <= 12%
● Median CV coverage < 0.75
● Coding coverage > 40X

RNA-Seq:GS sample-level matching was conducted using Somalier (Pedersen et al, 2020) inputting genomic variant call format files (gVCFs) for DNA and BAMS for RNA-Seq. Somalier was run with default settings including min-AB set to 0.2 and relatedness cut-off of 0.9 as recommended by Somalier for RNA-Seq.

Additionally, 134 individuals that did not have at least 30 million “mapped unique reads” according to RNA-SeQC2 were excluded from analysis, giving a final analysis set of RNA-Seq from 5412 individuals, all of which were included in the v18 100kGP data release.

### Identification of gene expression and splicing outliers

To identify gene expression and splicing outliers, OUTRIDER (Brechtmann et al, 2018) and FRASER2 (Scheller et al, 2023), respectively, were run via the DROP pipeline (Yepez et al, 2021) on batches of 500 samples at once for computational feasibility (a small number of individuals were included in multiple runs to enable complete batches of 500). The 11 batches were determined by the total number of uniquely mapped properly paired reads, such that samples with similar read counts were included in the same runs to minimise the impact differing read counts may have. Significance thresholds followed the DROP documentation’s default values. Splicing outliers were deemed “significant” if FRASER2 gave a difference in the percent spliced in (deltaPSI, a measure of difference in splice site usage) >= 0.1 or <= -0.1 and a false discovery rate (FDR) adjusted p-value < 0.1. Expression outliers were deemed “significant” if OUTRIDER gave an FDR adjusted p-value < 0.05. In some instances, unadjusted/raw p-values were also assessed to establish whether a given gene or event had been detected but had not passed correction for multiple testing.

Individuals with high numbers of outlier events from FRASER2 and OUTRIDER were identified as event count outliers (outlier event counts > mean + 2 standard deviations). To seek the potential underlying cause, difference in RNA-SeQC2 generated metrics were tested between these event count outliers and the rest of the group using wilcox.test in RStudio, with FDR adjusted p-values. Boxplots were generated using ggplot2 to visualise the differences in the top 10 most significantly different metrics for FRASER2 and OUTRIDER.

### Identification of known diagnoses identified through genome only analysis to act as positive controls

Although most of the sequenced individuals were undiagnosed at the time of selection, at the time of analysis, some did have confirmed or suspected diagnostic variants that could be used as positive controls to establish what the pipeline would detect.

Using information from Exit Questionnaires (see Web Resources) contained in the LabKey database, 368 cases were identified where the “solved” status had been recorded as “yes” or “partially”. RNA-Seq data from exit questionnaire confirmed diagnostic variants were reviewed in IGV to establish whether the impact of the variant on splicing could be observed in the RNA-Seq data and would therefore be potentially detectable by the outlier detection tools.

### Exploring events in ClinGen haploinsufficient genes

To enrich for RNA effects that are most likely to be disease relevant, we filtered the set of high-confidence gene expression and splicing outliers for genes robustly linked to disease via a haploinsufficient (HI) mechanism. A list of 376 genes coded as HI=3 was downloaded from the ClinGen resource (see Web Resources, accessed 9^th^ April 2024, **Table S1**).

FRASER2 outliers located in ClinGen HI genes and within PanelApp gene panels relevant for each individual were manually reviewed to assess: a) technical validity (i.e. whether the outlier is a genuine splicing disruption) using IGV, and b) phenotypic concordance, comparing the participant’s HPO terms and disease classification and the phenotype associated with loss-of-function of that gene as reported in OMIM. Genetic variants underlying the outlier events were identified from assessment of RNA-Seq and GS sequencing reads in IGV, and/or from GS VCFs. Plausible candidates were subsequently reviewed by a dedicated clinical curation team (Genomics England Clinical Research Interface), with credible candidates reported to the initial referral clinical teams.

### Structural variants impacting gene expression and splicing

We considered that a subset of expression and splicing outlier results may be due to the presence of rare structural variants. RNA outliers were therefore filtered for those where the same individual also harboured a rare structural variant affecting the same gene. To do this we used a merged set of variant calls from Manta and Canvas algorithms (Chen et al, 2016; Roller et al, 2016). These structural variants (SVs) were entered into a mysql database and clustered using an 80% overlap threshold, as described previously (Yu et al, 2022). Only deletions, duplications and inversions were considered due to the difficulties of interpreting insertions and “breakend” calls at scale. However, for genes linked to a highly specific phenotype and where the participant’s clinical information matched that condition, read alignments were reviewed manually in a small number of cases.

For Case Study 7, long read sequencing was used to further investigate the nature of the SV identified. A nanopore sequencing library was prepared from 3 ug of genomic DNA following the manufactures protocol for kit LSK114 (Oxford Nanopore Technologies). The library was sequenced on a P48 PromethION using a single R10 flowcell, with nuclease washing and library re-loading taking place at 24- and 48-hours respectively. Basecalling and alignment to the human reference genome was performed using Dorado v.0.7.3 (super high accuracy model). The sequencing run yielded 106.65 Gb of sequence data (17.17 million reads) with an N50 of 14.79 kb. Long-read RNA sequencing was performed following the manufacturers protocol for kit SQK-PCS114 and run on a single R10 flowcell using a P48 PromethION. Dorado v.0.7.3 was used to basecall (high accuracy model) and align individuals reads to the human reference genome using the “splice” preset. The sequencing run yielded 128.83 Gb of sequence data (107.51 million reads) with an N50 of 1.29 kb.

### Exomiser prioritisation of diagnostic candidates

Exomiser 12.1.0 was used with the 2102 database release and default settings for identification of rare, segregating, predicted deleterious, coding variants with the additional retention of intronic variants and SpliceAI scoring of SNVs using the spliceai_scores.raw.snv.hg19.vcf and spliceai_scores.raw.snv.hg38.vcf files downloaded (see Web Resources). This version of Exomiser was run on multi-sample GS files for the 5412 samples that had undergone RNA-Seq.

A set of potential diagnoses was obtained from the Exomiser output by identifying all candidates where:

- The affected gene was identified as a gene expression or splicing outlier
- The affected gene was classified as a green gene on a relevant PanelApp gene panel or had an Exomiser human phenotype score > 0.6 indicating strong similarity between the patient’s phenotype and reference annotations from OMIM or Orphanet diseases associated with the same gene (and the correct mode of inheritance (MOI))
- The variant had a SpliceAI score > 0.2
- The variant was in the coding region or within the first or last 50bp of an intron

After review, the 74 potential diagnoses were classified into:

- 35 known diagnoses with 28 involving at least one canonical splice variant
- 39 candidate diagnoses with 20 involving at least one canonical splice variant

These 39 candidate diagnoses were classified by evaluating further evidence such as the predicted ACMG classification from a reanalysis using Exomiser 14.1.0 and ClinVar annotations. Phenotypic relevance was assessed by reviewing Human Phenotype Ontology (HPO) terms (Gargano et al, 2024) recorded for each proband and comparing them with the hallmark clinical features of disorders associated with the implicated gene. Finally, RNA-Seq data were manually inspected in IGV to evaluate the presence and nature of splicing alterations, and to characterise their consequences in relation to each candidate variant. SpliceAI (Jaganathan et al, 2019) and Pangolin (Zeng & Li, 2022) scores were assessed to aid interpretation. Representative examples of these candidates are given in Results, with further detail in Case Studies 8, 9, and 10.

To identify additional splice-altering variants that may lie outside canonical splice sites and beyond the ±50 bp boundaries, an extended analysis was conducted.

A total of 2,641 variants were extracted from the Exomiser output by identifying all variants that met the following criteria:

- The affected gene was identified as a gene expression or splicing outlier
- The variant had a SpliceAI score > 0.2

Variants located at canonical splice sites and those from individuals with a confirmed diagnosis were excluded, leaving 1,627 variants for further evaluation.

Phenotypic prioritisation was then applied, retaining only variants that occurred in a gene classified as a green PanelApp gene on the recruited disease panel or received an Exomiser human phenotype score > 0.6 consistent with the correct mode of inheritance. This filtering step resulted in 88 candidate variants.

Of these, 39 fell within the 50 bp splice region and corresponded to those described above. The remaining 49 variants, which were located beyond the 50bp, were assessed using the same strategy: Exomiser automated predicted ACMG classification, phenotypic relevance based on HPO terms, and manual inspection of RNA-Seq data in IGV. A representative example of these candidates is shown in Example 11.

## Results

### Characteristics of the sequenced cohort

The final analysis cohort comprised 5,412 individuals for whom the whole-blood RNA-Seq data passed QC (see Methods). Most participants (n=5,401) were recruited to the 100kGP as probands, with 11 individuals recruited as affected relatives. Age at recruitment showed a bimodal distribution, with peaks at 5-9 years and 45-49 years, and a median of 27 years (range 0-93, **Figure 1a**). There were 2,652 participants whose declared phenotypic sex was recorded as being female (49%), while the remaining 2760 were male (51%). The inferred karyotypic sex was concordant with the reported phenotypic sex for 5390 individuals (99.65%). For 17 individuals there was either a mismatch between their phenotypic and inferred karyotypic sex, or a non-XX/XY inferred karyotypic sex (including XO, XXY, XYY, XXXY). For the remaining 5 individuals, the inferred karyotypic sex was missing/inconclusive. Of the 4944 individuals whose ancestry could be inferred from their genetic data (91.35%), 4314 participants (87.26%) were of European ancestry, with much lower representation of other individual ancestries (**Figure 1b-c**).

**Figure 1:**
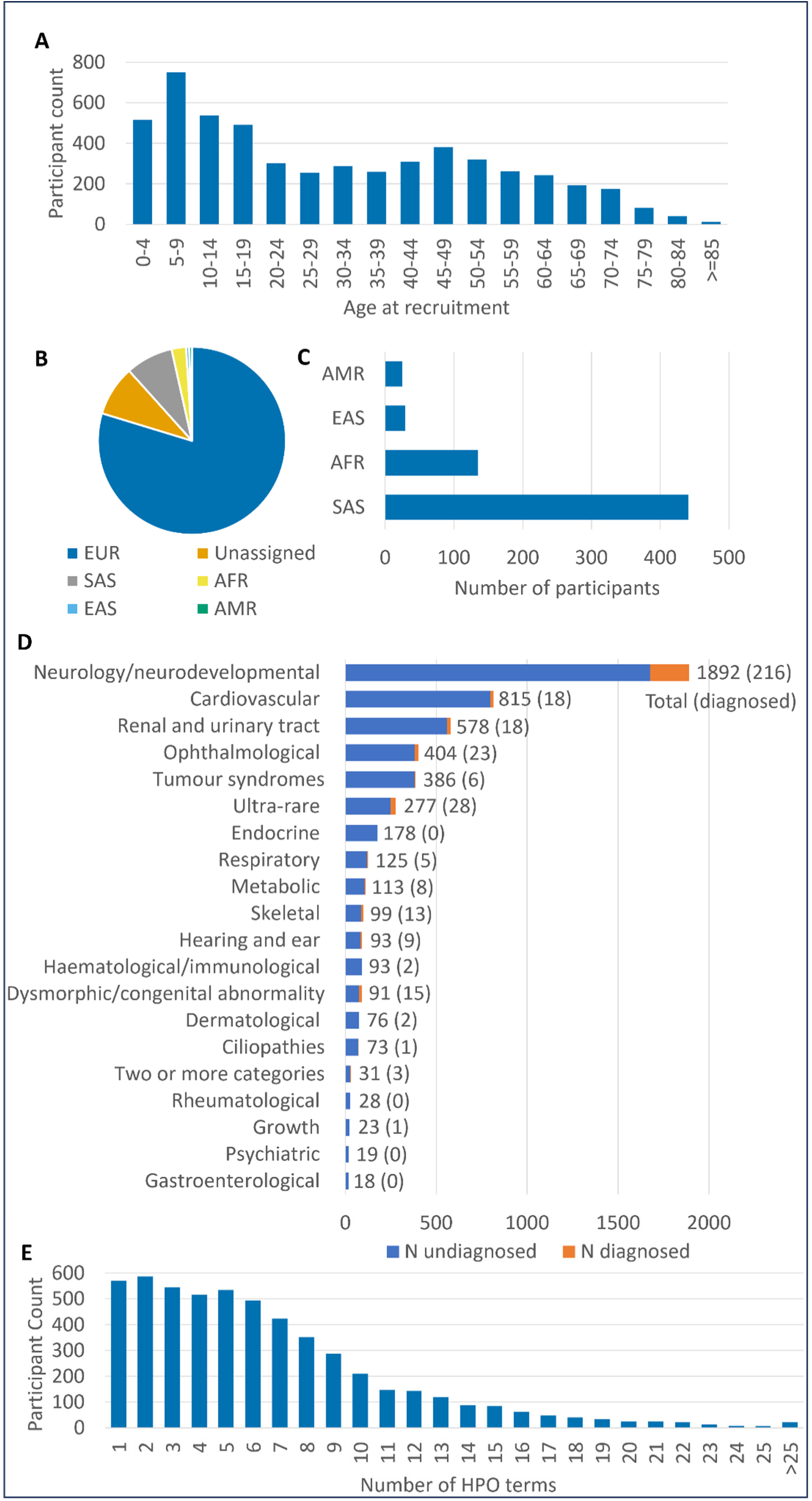
Summary of demographics of 5,412 individuals in RNA-Seq cohort. A) Participant age at time of recruitment/sample collection in years. B) Genetically-inferred ancestry shows the majority of the cohort are of European ancestry. EUR = European, SAS = South Asian, EAS = East Asian, AFR = African, AMR = Admixed American ancestry. C) Breakdown of genetically inferred ancestry with unassigned and EUR individuals removed for clarity. D) Number of individuals recruited under different “normalised disease groups”, including number with a confirmed genetic diagnosis at time of analysis. E) Number of Human Phenotype Ontology (HPO) terms marked as present in participants at recruitment.

At recruitment, participants were assigned to one of 19 “Normalised Disease Groups” (**Figure 1d**). The largest group was “Neurology and neurodevelopmental disorders” (n=1,892, 34.96%), followed by “Cardiovascular disorders” (n=815, 15.06%) and “Renal and urinary tract disorders” (n=578, 10.68%). 31 individuals (0.57%) were recruited under more than one disease classification. At time of analysis, 368/5412 (6.80%) individuals in the cohort had a confirmed genetic diagnosis. Diagnostic rates varied by disorder type (**Figure 1d**), with rates above 10% for “Dysmorphic and congenital abnormality syndromes” (15/91, 16.48%), “Skeletal disorders” (13/99, 13.13%), “Neurology and neurodevelopmental disorders” (216/1892, 11.42%) and “Ultra-rare disorders” (28/277, 10.11%).

In addition to the high-level disease classification, HPO terms were also recorded for participants. The number of HPO terms reported as present for individuals was highly variable (median = 5, range 1-38, **Figure 1e**). Across the cohort, 2,350 individuals (43.4%) had a single PanelApp gene panel assigned to them at the time of first diagnostic analysis, while the median number of gene panels assigned was 2 (range 0-16). Nine individuals had no relevant PanelApp panels recorded in LabKey. The median number of genes within these relevant panels (i.e. the number of genes assessed per participant) was 235 (range 0-4087). For 29 individuals, there were no relevant genes as they either did not have any gene panels assigned (n=9), or their assigned gene panels contained no diagnostic grade (“green”) genes (n=20, including the severe familial anorexia, juvenile dermatomyositis, PHACE(S) syndrome panels).

### Characteristics of the sequencing

A median of 110.64 million mapped unique reads per individual were generated (range 30.21-403.12, **Figure S1a**). Most of these reads mapped to intronic regions of the genome (median 62.0%) as has been previously reported for rRNA depleted RNA-Seq, likely due to capture of immature mRNA (Zhao et al, 2018). A median of 28.8% mapped to exons, with smaller proportions of intergenic (5.1%) and ambiguously located reads (mapping to multiple genes, or intronic and exonic regions, 4.0%, **Figure S1b**). The best captured type of genes were protein coding genes, with around 51.7% (10,314/19,965) having a median transcripts per million (TPM) expression level ≥ 5. Around 6.9% (1170/16,849) of long non-coding RNAs (lncRNAs) had a median TPM ≥ 5 across the cohort, with much smaller proportions of other RNA species well represented in the data (4.0% snoRNAs (38/942), 1.4% snRNAs (27/1901), 16.3% scaRNAs (8/49), and 0.1% of miRNAs (1/1881), **Figure S1c**).

Across protein coding genes, a greater proportion of genes associated with disorders (i.e. “green” PanelApp genes) had a median level of expression of TPM ≥ 5 than did those not associated with a disease (51.7% (10,314/19,965) across all genes, 59.6% (2295/3849) across all PanelApp genes (chi square p=2.2x10^-16^), **Figure 2a**). For 95.2% of the PanelApp genes with median expression TPM ≥ 5 (2184/2295), at least 80% of the cohort had individual TPMs ≥ 5, indicating relatively consistent expression of these genes across the cohort. The proportions of genes in different panels with TPM ≥ 5 varied from 32% for the congenital myopathy panel to 92% for the congenital disorders of glycosylation panel (**Figure 2b**).

**Figure 2:**
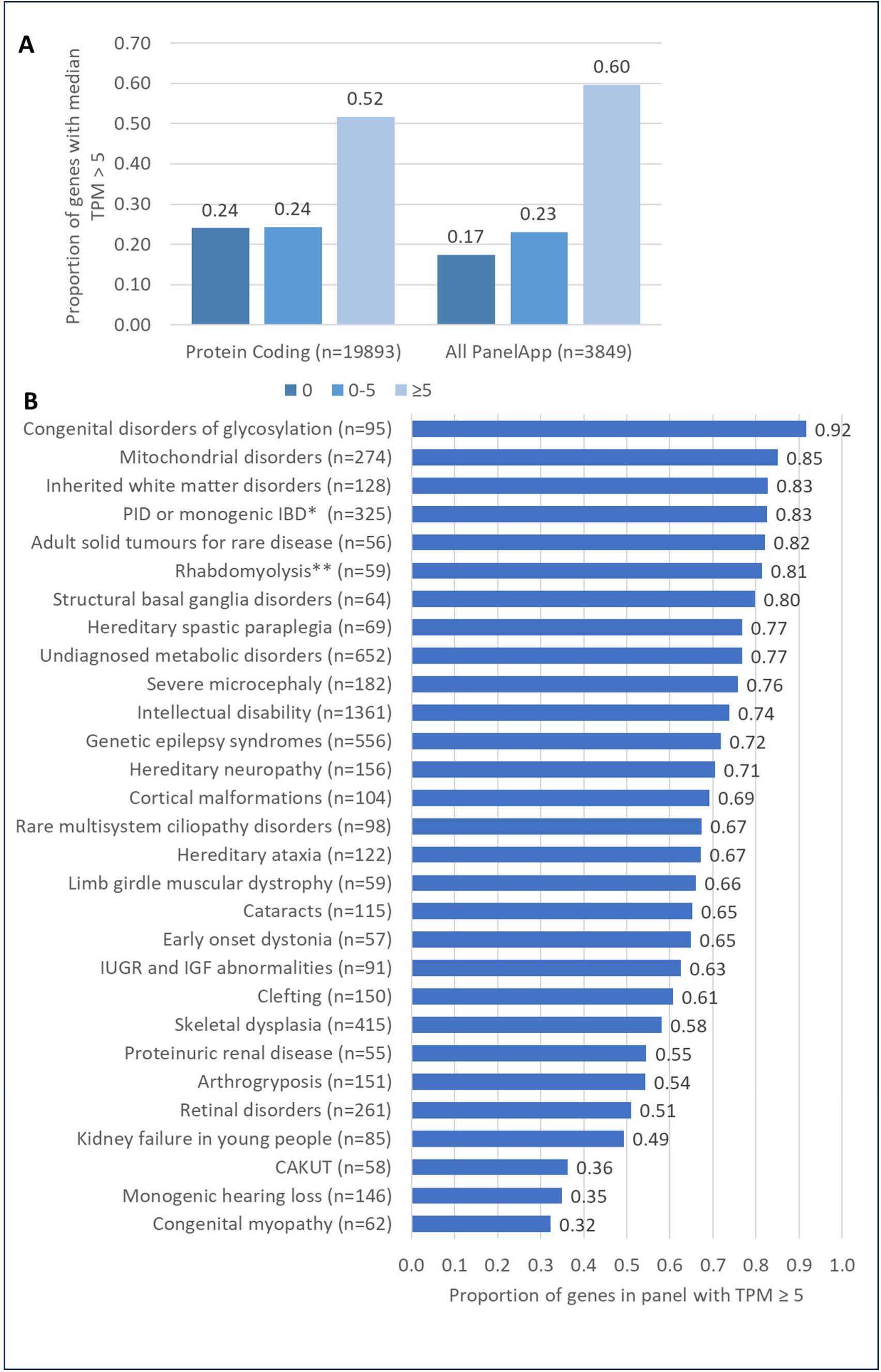
Proportions of genes captured by the RNA-Seq data. A) Proportion of all genes and all PanelApp genes with median expression 0, 0-5 and ≥ 5 TPM. B) Proportion of genes in different PanelApp panels with TPMs ≥ 5. Plot showing all gene panels with at least 50 “green” (high confidence) genes which were applied to at least 50 of the RNA-Seq cohort. *Primary immunodeficiency or monogenic inflammatory bowel disease. **Rhabdomyolysis and metabolic muscle disorders.

### Identification of outliers and outlier characteristics

Gene expression and splicing outliers were identified using OUTRIDER and FRASER2 respectively, to identify potentially disease-associated events. Across the cohort, 36,239 gene expression outliers in 10,478 unique genes and 42,258 splicing outliers in 10,902 unique genes were detected (**Table 1, Figure S2**).

**Table 1:**
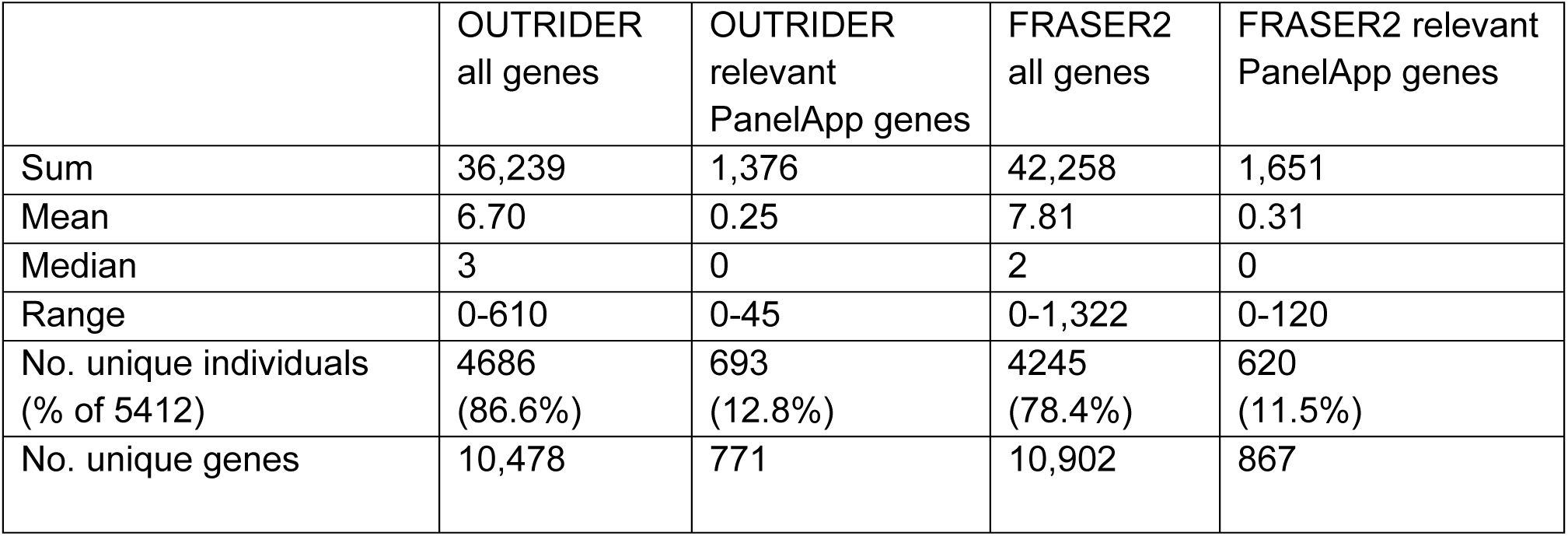
Summary of the number of expression and splicing outlier events identified across all genes and in genes in relevant PanelApp gene panels.

|  | OUTRIDER<br>all genes | OUTRIDER<br>relevant<br>PanelApp genes | FRASER2<br>all genes | FRASER2 relevant<br>PanelApp genes |
| --- | --- | --- | --- | --- |
| Sum | 36,239 | 1,376 | 42,258 | 1,651 |
| Mean | 6.70 | 0.25 | 7.81 | 0.31 |
| Median | 3 | 0 | 2 | 0 |
| Range | 0-610 | 0-45 | 0-1,322 | 0-120 |
| No. unique individuals<br>(% of 5412) | 4686<br>(86.6%) | 693<br>(12.8%) | 4245<br>(78.4%) | 620<br>(11.5%) |
| No. unique genes | 10,478 | 771 | 10,902 | 867 |

To focus on the events most likely to be relevant to the participants’ reported phenotypes, we restricted analyses to genes included in PanelApp panels flagged as relevant for each individual by the Genomics England pipeline at time of recruitment. Considering just these potentially diagnostically relevant genes, a total of 1,376 gene expression outliers (771 unique genes) and 1,651 splicing outliers (867 unique genes) were observed when adjusting significance thresholds to account for transcriptome-wide significance (**Table 1**). 1,091 individuals had at least one gene expression or splicing outlier in a gene from a relevant gene panel (20.2% of the cohort).

A number of individuals were identified as event count outliers with high numbers of splicing (n=74) and/or expression (n=66) outlier events identified (>mean+2 standard deviations, 15 individuals were event count outliers for both splicing and expression). These individuals were found to have significantly different RNA-SeQC2 (Graubert et al, 2021) derived scores across a variety of metrics (**Table S2, Figure S3**). The most significantly different metrics for both OUTRIDER and FRASER2 were mapped_unique_reads (number of aligned reads without duplicate flags) and genes_detected (the number of genes with at least five unambiguously mapped reads), with fewer mapped unique reads and fewer genes detected in event count outliers relative to the rest of the cohort.

There was limited overlap in the genes identified as both splicing and expression outlier events (i.e. the same gene identified in the same individual as an outlier by both OUTRIDER and FRASER2). Just 3.0% (1,250/42,258) of splicing outlier events occurred in genes which were also significant expression outliers for that individual. 17.8% (7534/42,258) of splicing outliers resulted in a difference in expression at nominal significance, but which was not significant after FDR correction for multiple testing. The percentage overlapping was slightly higher for events in genes in relevant PanelApp panels, with 4.7% (77/1,651) of splicing outliers also resulting in FDR corrected significant expression outlier events, and 23.2% (383/1,651) resulting in a nominally significant expression outlier event.

### Sensitivity of the RNA-Seq outlier approach to confirm known diagnoses identified through genome only analysis

Although most individuals who underwent RNA-Seq were undiagnosed at the time of sample selection, a small subset had or later received, confirmed genetic diagnoses through the analysis of GS datasets. Of the 368 individuals with genetic diagnoses from GS alone, 47 were identified with splicing disruptions through manual inspection of RNA-Seq reads in IGV (**Table S3**). Automated detection using FRASER2 identified 23 of these splicing events (48.9%) at transcriptome-wide significance after FDR correction. All 47 events were identified using FRASER2’s event-based uncorrected p-values, but this substantially reduced specificity, increasing the number of candidate splicing outliers in relevant gene panels for the 47 individuals from 31 at genome-wide significance to 17,335 at nominal significance.

The events which reached transcriptome-wide significance had higher absolute deltaPSIs than did those only at nominal significance (median 0.48 for significant events relative to 0.31 for nominal group, t-test p-value=0.000993), and were in genes with higher TPM values across the cohort (median 126.1 for significant events relative to 42.9 for nominal group, t-test p-value=0.00993). There was no difference in the individual’s number of mapped unique reads, the direction of effect of the splicing event (negative or positive deltaPSI), or whether they also were identified as outliers by OUTRIDER (**Table S4, Figure S4**). Overall, just under half (48.9%) of these diagnostic, manually confirmed splicing abnormalities were detected by FRASER2 at transcriptome wide significance.

### Identifying potential diagnostic candidates

With over 3,000 significant outlier events in disease relevant genes identified and a significant amount of manual review required to fully assess the technical validity and potential clinical relevance of these candidates, several strategies were employed to prioritise the most probable diagnostic candidates. These strategies included leveraging information on the genes in the genome most likely to harbour dominant pathogenic genetic variants that may explain the detected outlier events. Specifically, we assessed splicing outliers in ClinGen HI genes in relevant gene panels for each individual, overlap of outliers with structural variation, and utilising Exomiser (which incorporates many different types of data including phenotypic relevance) to identify and prioritise genetic variation that likely explained disease relevant outlier events.

### Review of splicing outliers in ClinGen haploinsufficient genes in relevant PanelApp panels identifies likely diagnostic variants

There were 200 FRASER2 splicing outliers in 129 individuals without prior diagnoses which fell in ClinGen HI genes and which were on gene panels relevant for that individual. Manual inspection of all events using IGV gave 78 genuine, clearly visible splicing aberrations (39% of 200 events), and 70 events (35%) which failed manual verification (i.e. no evidence of abnormal splicing was observed, or abnormal splicing was also observed at comparable levels in individuals without this event identified as an outlier). The remaining 52 events (26%) could not be reliably classified (often these were regions with low coverage where some evidence of abnormal splicing could be observed but not enough for confident classification). In general, the events which were manually confirmed had higher sequencing depth (mapped unique reads, t-test p=0.01) and lower p-values (t-test p=0.01), but there was no difference between the magnitude of the events (absolute deltaPSI, t-test p=0.10) or the proportion of negative deltaPSI scores (chi square p=0.08) (**Table S5, Figure S5**). There was a difference in the proportion of events verified between the different types of event called by FRASER2 (Fisher’s exact p=2.09x10^-05^, e.g. over half of “exonTruncation” events and “exonSkipping” events were manually verified, while only 17% of “annotatedIntron_increasedUsage” were, **Table S5**), which may reflect difficulties in the manual verification process of certain event types where subtle changes in annotated events are observed, rather than the usage of a novel splice site or splice site combination. A larger percentage of the splicing events which passed manual verification also resulted in expression outlier events (12.8% (10/78)) than did those which failed validation (2.9% (2/70)).

While full investigation of all potential diagnostic events is still ongoing, here we present several examples which capture the range of different types of variant and splicing impacts identified in the cohort (**Table 2**, further information (FRASER2/OUTRIDER outlier data, gnomAD minor allele frequencies, SpliceAI scores) in **Table S6**, extended case studies in **Supplemental Case Studies**, SpliceAI and Pangolin prediction scores in **Figure S6**).

**Table 2:** Summary variant, outlier and phenotype information for featured Case Studies

| Case study no. | Variant (GRCh38 coordinates, ENSG) | Gene | Normalised Specific Disease | FRASER2 outlier* | OUTRIDER outlier* | Impact | Mechanism |
| --- | --- | --- | --- | --- | --- | --- | --- |
| 1 | del(chr3:41236871-41238422) | *CTNNB1* | Intellectual disability | Yes | Nominal | Exon skipping | Deletion of "skipped" exon |
| 2 | chr6:79060476:A:C,<br>ENST00000275034.5:c.439+2T>G | *PHIP* | Intellectual disability | Yes | Nominal | Exon skipping | Splice donor disruption |
| 3 | chr1:224404379:A:C,<br>ENST00000414423.9:c.1599+51T>G | *WDR26* | Intellectual disability | Yes | No | Partial intron retention | Cryptic donor activation |
| 4 | chr12:49042352:T:G,<br>ENST00000301067.12: c.5868-22A>C | *KMT2D* | Kabuki syndrome | Yes | Yes | Partial intron retention | Branchpoint disruption |
| 5 | chr2:32128139:C>T,<br>ENST00000315285.9:c.1174-269C>T | *SPAST* | Hereditary Spastic Paraplegia | Nominal | Yes | Pseudoexon creation | Cryptic site activation |
| 6 | del(chr1:91907874-92835348) | *RPL5* | Congenital anaemias | Nominal | Yes | Decreased expression | Deletion of first 4 exons |
| 7 | APC intron 10 retrotransposition | *APC* | Multiple bowel polyps | Yes | Yes | Increased expression | Intronic retrotransposition |
| 8 | chr2:47813234:C:T,<br>ENST00000403359.7:c.2227G>A | *FBXO11* | Intellectual disability | Nominal | Yes | Exon skipping, intron retention | Splice donor disruption |
| 9 | chr11:72234615:G:A,<br>ENST00000298229.7:c.2415G>A | *INPPL1* | Chondrodysplasia punctata | Yes | Yes | Partial intron retention | Splice donor disruption |
| 10 | chr2:71681110:G:A,<br>ENST00000410020.8:c.6173G>A | *DYSF* | Limb-girdle muscular dystrophy | Yes | Nominal | Exon skipping, partial intron retention | Splice donor disruption |
| 11 | chr2:25240802:T:C,<br>ENST00000321117.9:c.2083-72A>G | *DNMT3A* | Intellectual disability | Yes | No | Pseudoexon creation | Cryptic donor activation |

An exon skipping event was identified in *CTNNB1* (FRASER2 deltaPSI=-0.32, gene level adjusted p-value=3.82x10^-5^, **Figure 3ai**, **Case Study 1**) in a proband with intellectual disability, microcephaly and spasticity, consistent with CTNNB1 associated neurodevelopmental disorder with spastic diplegia and visual defects (MIM:615075). Manual inspection of the individuals’ DNA sequencing reads in IGV revealed a short, 1.55kb deletion spanning the “skipped” exon 14 (Figure 3aii). This had been identified by the Manta structural variant caller (Chen et al, 2016) but was filtered out of the analysis set as a low quality CNV call, likely due to low coverage.

Although *CTNNB1* was not identified as a significant expression outlier, a modest reduction in abundance was observed (OUTRIDER FC=0.88, uncorrected p-value=0.00037).

All genes monoallelic, except *INPPL1* which had compound heterozygous variants (frameshift 1bp deletion in addition to splice donor variant) and *DYSF* where the variant was homozygous. *Full details (deltaPSI, fold change, adjusted and raw p-values) given in **Table S6**.

A splicing outlier event in *PHIP* was identified (skipping of exon 6, deltaPSI=0.48, adjusted p-value=0.009, **Figure 3b, Case Study 2**) in an individual with intellectual disability, developmental delay and prominent forehead, consistent with Chung-Jansen syndrome (MIM:617991). The splicing disruption is thought to result from a variant near the splice donor site (chr6:79060476:A:C (GRCh38)) which had previously been assessed for this participant and was deemed to be a variant of uncertain significance (VUS). The RNA-Seq provides evidence of functional impact, and while the exon skipping is likely to be in-frame (removal of 34 amino acids), a nearby likely pathogenic variant in ClinVar (VCV002664249.1) affecting the same splice site strengthens evidence of pathogenicity.

**Figure 3.**
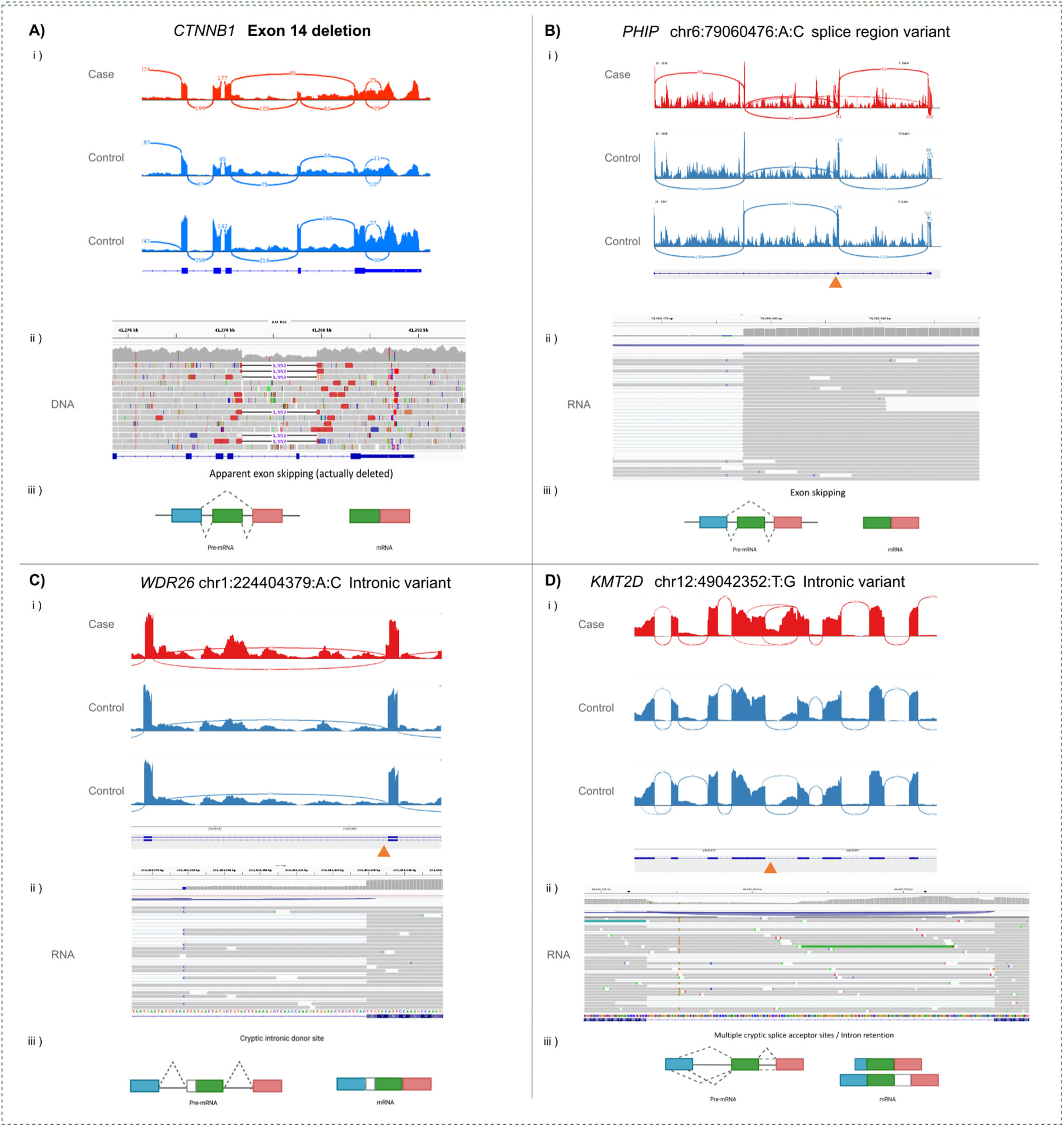
– Example candidate diagnoses identified through assessment of FRASER2 splicing outliers in ClinGen haploinsufficient genes. Each panel contains gene and variant information, i) an IGV generated Sashimi plot showing the nature of the splicing disruption in the individual with the variant relative to RNA-Seq from two other individuals without that variant, ii) an IGV screenshot of sequencing reads (DNA or RNA specified) showing the variant underlying the disruption, and iii) a schematic diagram summarising the observed impact(s) on splicing.

In *WDR26*, an exon elongation/partial intron retention event was identified (deltaPSI=0.18, adjusted p-value=0.02, **Figure 3c, Case Study 3**) in an individual with intellectual disability, developmental delay and seizures, consistent with Skraban-Deardorff syndrome (MIM:617616). This is thought to result from an intronic variant (chr1:224404379:A:C (GRCh38)) generating a cryptic splice donor site which can out compete the real exon 8 splice donor site. This would lead to the inclusion of 51bp intronic sequence in transcripts, which includes a termination codon. While this is a *de novo* variant in a relevant disease gene, its intronic location meant it was not prioritised from GS alone.

In an individual recruited with suspected Kabuki syndrome (MIM:147920), a partial intron retention event was identified in *KMT2D* (deltaPSI=-0.37, adjusted p-value=1.22x10^-9^, **Figure 3d, Case Study 4**), a gene consistent with their phenotype. Complex splicing disruptions, including intron retention and the use of multiple cryptic sites were observed. This is likely due to a variant 22bp upstream of the exon 29 splice acceptor site (chr12:49042352:T:G (GRCh38)) disrupting the splicing branchpoint, which is predicted by LaBranchoR (Paggi & Bejerano, 2018) to be deleterious to the branchpoint’s function.

An additional splice-disrupting variant in *SPAST* (MIM:182601) was identified through analysis of OUTRIDER gene expression outliers (FC=0.75, adjusted p-value=0.00026, **Case Study 5, Figure S7**) in an individual with hereditary spastic paraplegia (HSP). The region was flagged as a splicing outlier but did not withstand correction for multiple testing (deltaPSI=-0.3, adjusted p-value=0.19427). A deep intronic variant (chr2:32128139:C>T (GRCh38)) was observed 268bp from the exon 9 splice acceptor site, leading to the creation of a pseudoexon including a premature termination codon. The same variant was also observed in three other individuals recruited with HSP, along with one affected individual’s father who had not been assessed for the condition.

### Intersection of outliers with structural variants highlights diagnostic candidates

OUTRIDER expression outliers contain both up and down regulated genes. Transcriptome-wide, the ratio of up and down regulated genes is almost 1:1 (n upregulated / n downregulated = 1.01). However, across the haploinsufficient (HI=3) gene-set, there was a clear skew against downregulation (n upregulated / n downregulated = 1.40). We hypothesised that for many of the OUTRIDER gene expression outliers, the result could be due to copy number or other types of structural variants. This idea was explored by assessing the 766 OUTRIDER high-confidence gene expression outliers in the 376 haploinsufficient genes. Intersection of expression outlier calls with LOSS and INVERSION calls using the SVRare database identified 37 participants where the gene disrupted by the structural variant matched the gene shown to be an expression outlier in the RNA-Seq data. Following manual review of these, we found that in 19/37 (51%) cases, the SV call appeared to be genuine, and could help explain the RNA-Seq result. In contrast, there were 1,059 outliers across the 376 genes using FRASER2 splicing outlier results, and of these only 19 individuals harboured a rare SV in the candidate gene of interest.

RNA-Seq results for six participants harbouring genomic inversions have been reported previously (Pagnamenta et al, 2024) and together highlight the utility of RNA-Seq to support inversion variant pathogenicity. The most notable example was a 764kb inversion disrupting the 5’-UTR of *DYRK1A*. The reduced expression (FC=0.57, adjusted p-value=3.7x10*^-27^*) helped confirm that exon 1 (which lacks protein coding content) is critical for normal gene expression. Another participant with intellectual disability due to a deletion-inversion involving *SRRM2* and published as P1 in Pagnamenta *et al* (Pagnamenta et al, 2023) was shown to exhibit significantly reduced *SRRM2* expression (FC=0.41, adjusted p-value=1.24x10^-31^).

Here, using an RNA-first approach we identified another case with disruption of a 5’-UTR exon, this time involving *ANKRD11.* There was evidence of significantly reduced *ANKRD11* expression (FC=0.54, adjusted p-value=1.21x10^-33^) for this individual, likely due to a 5.6kb deletion on 16q24.3 (**Figure S8a**). Deletions of this region are reported in the literature to impact normal gene expression (Borja et al, 2023). Loss of *ANKRD11* is linked to KBG syndrome (MIM: <u>148050</u>), characterised by craniofacial abnormalities, short stature and developmental delay.

This proband was recruited to the 100kGP under the Intellectual Disability classification, with delayed speech and language, delayed fine and gross motor development and global developmental delay.

In an individual recruited with congenital anaemia, OUTIRDER identified a gene expression outlier in *RPL5* (FC=0.56, adjusted p-value=2.82x10^-12^, **Figure S9a, Case Study 6**), causal of Diamond-Blackfan anaemia 6 (MIM: 612561). This intersected with a large, 927kb deletion affecting the first 4 exons of the 8-exon gene (approx. coordinates chr1:91907874-92835348 (GRCh38)). Three of seven additional protein coding genes in the deleted region were also identified as outliers by OUTRIDER (**Figure S9b**).

For several individuals, the dysregulated gene identified by OUTRIDER analysis was linked to a highly specific condition, closely matching the clinical information available for the proband being assessed. For such cases, read-alignments were viewed manually, regardless of whether an SV had been identified. In an individual recruited with multiple bowel polyps and with significant family history, *APC* was identified as being significantly upregulated (FC=1.58, adjusted p-value=6.91x10^-25^, **Case Study 7**). Much of the signal appeared to localise to intron 10, and although no SVs were identified here, evidence of split-read pairs was observed in both the GS and RNA-Seq data (**Figure S10**). Using nanopore long-read DNA sequencing (**Figure S11**) with Sanger sequencing confirmation of breakpoints (**Figure S12)**, this was resolved as an SVA type D retrotransposition event, which was also present in 4/4 of the proband’s affected relatives (**Figure S13**).

### Exomiser highlights candidate novel diagnoses

Excluding known diagnoses, the 39 variants flagged by Exomiser (as described in the Methods) were assessed based on the strength of the combined genetic, transcriptomic, SpliceAI scores and clinical evidence. 16 candidates showed robust support, with clear splice-altering effects observed in the RNA-Seq data and strong consistency with the expected disease phenotype and mode of inheritance; 21 candidates were classified as highly promising but required additional clinical data, segregation information, or phenotypic clarification; 2 candidates were excluded due to insufficient phenotypic overlap with the gene’s associated disease under the observed mode of inheritance. Representative examples of the compelling candidates are discussed below.

A missense variant (chr2:47813234:C:T, (GRCh38)) was identified in an individual recruited with intellectual disability in *FBXO11* (**Figure 4a, Case Study 8**), associated with *FBXO11*-related autosomal dominant intellectual developmental disorder with dysmorphic facies and behavioural abnormalities (MIM 618089). This was the top ranked Exomiser variant for this individual (Exomiser combined score=0.948) and was flagged as a gene expression outlier (FC=1.32, adjusted p-value=3.21x10^-13^). Inspection of the RNA-Seq data showed the variant, which falls in the conserved last base of the exon, causes skipping of exon 18 and retention of surrounding intronic sequence, which likely underlies the apparent overexpression of the gene. Despite the variant having a clear impact on splicing (**Figure 4ai**), it was not identified as a significant splicing outlier (FRASER2 deltaPSI=-0.45, adjusted p-value=0.179, raw p-value=2.45x10^-6^). The variant had already been assessed through the Genomics England diagnostic pipeline, and deemed to be a VUS, but evidence from the RNA-Seq supports reclassification of this variant as likely pathogenic.

**Figure 4.**
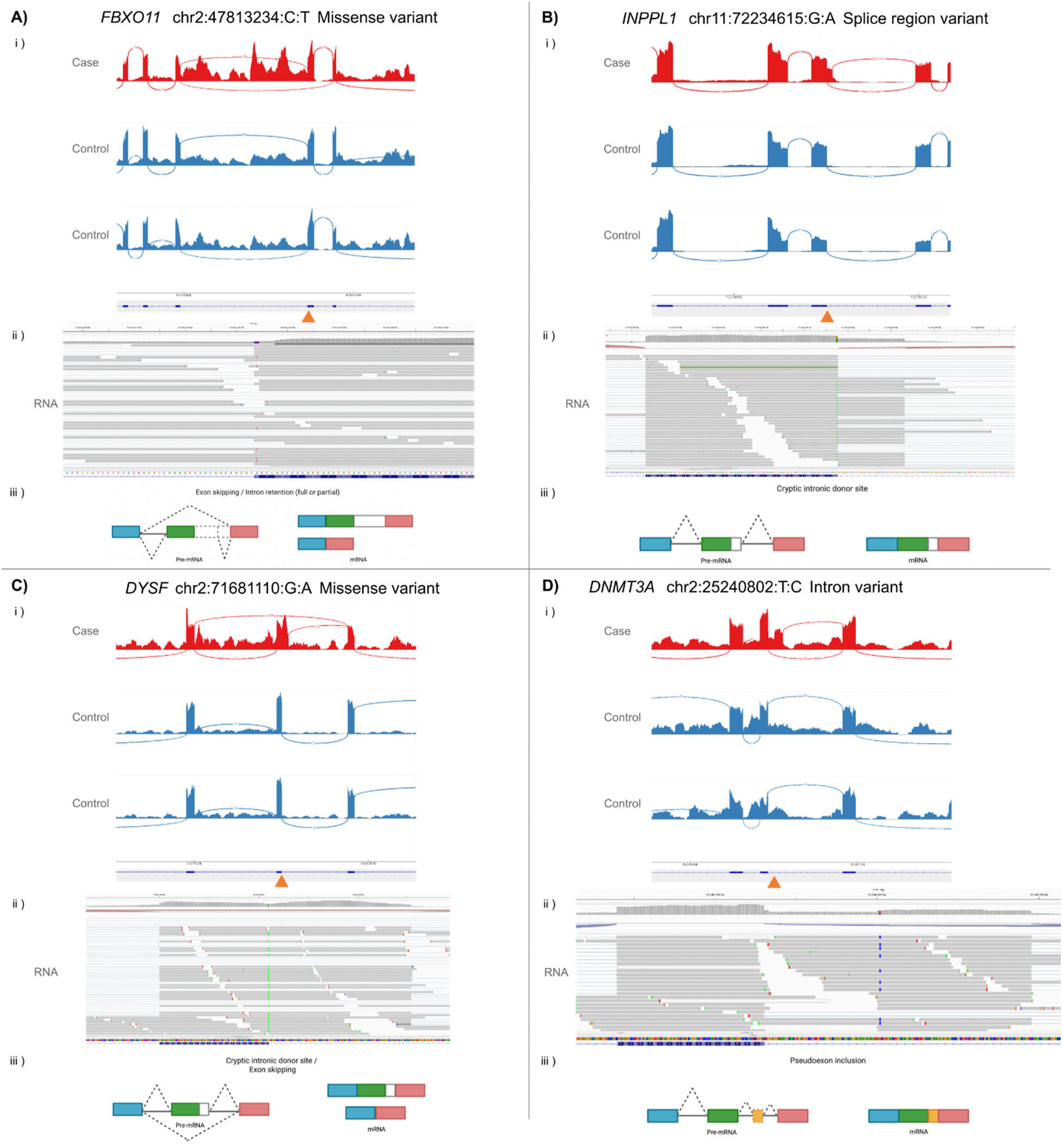
-. Example candidate diagnoses identified through assessment of splicing and gene expression outliers overlapping with Exomiser candidate variants. Each panel contains gene and variant information, i) an IGV generated Sashimi plot showing the nature of the splicing disruption, ii) an IGV screenshot of RNA sequencing reads showing the variant underlying the disruption, and iii) a schematic diagram summarising the observed impact(s) on splicing.

In an individual recruited under the chondrodysplasia punctata disease group, compound heterozygous variants were identified in *INPPL1* (Exomiser combined score=0.830), associated with *INPPL1*-realted opsismodysplasia (MIM:258480, **Case Study 9**). These were a synonymous variant in the last base of exon 21 (chr11:72234615:G:A (GRCh38) and a frameshift variant (1bp deletion, chr11:72237532:GC:G (GRCh38)). FRASER2 identified reduced usage of the normal splice donor site, with activation of a cryptic donor site 31bp away (deltaPSI=-0.61; adjusted p-value=2.85×10⁻⁶, **Figure 4b**), leading to the inclusion of 31bp of intronic sequence, leading to a frameshift effect. The gene was also identified by OUTRIDER with significantly reduced expression (FC=0.72, adjusted p-value=3.15×10⁻⁴). It is notable that there was a relatively modest impact on expression despite two apparent loss-of-function variants that would be expected to result in nonsense mediated decay. The variant pair had previously been assessed through the Genomics England diagnostic pipeline, with the synonymous variant being classified as a VUS. The demonstrated splicing disruption supports the reclassification of the variant as likely pathogenic.

In a proband recruited with limb-girdle muscular dystrophy, a partial intron retention event (deltaPSI=–0.92, adjusted p-value=1.30×10⁻⁴⁹, **Figure 4c, Case Study 10**) was identified in *DYSF*, associated with Muscular Dystrophy, Limb-Girdle, Autosomal Recessive 2 (LGMDR2, MIM:253601). Exomiser prioritised a homozygous missense variant (chr2:71681110:G:A (GRCh38), Exomiser combined score=0.637) in the last nucleotide of exon 54. The RNA-Seq data showed the use of a cryptic splice donor site, 143bp downstream of the normal splice site, as well as exon skipping of exon 54, both of which would be expected to have a frameshift impact.

Finally, after expanding the search area beyond 50bp from the intron:exon boundaries, Exomiser highlighted a variant (chr2:25240802:T:C (GRCh38), Exomiser combined score=0.823, **Case Study 11**) in *DNMT3A* in a proband recruited under the intellectual disability disease group, and with HPO terms consistent with Tatton-Brown-Rahman syndrome (TBRS, MIM:615879). FRASER2 identified a significant splicing outlier event (deltaPSI=–0.43; adjusted p-value=0.010778), which manual inspection revealed to be due to the use of a cryptic intronic donor site at the variant position (**Figure 4d**), creating a 93bp pseudoexon between exons 17 and 18. Although paternal DNA was not available to confirm *de novo* status, the variant was not maternally inherited, has a clear impact on splicing, and the gene matches the phenotype seen in the proband, making this a strong diagnostic candidate.

## Discussion

The use of RNA-Seq as a complementary approach to exome and genome sequencing in rare disease diagnostics has been rapidly expanding in recent years and applied across a diverse range of disorders. Here, we present the largest cohort to date leveraging RNA-Seq to improve diagnostic yield for rare disorders. Unlike most previous studies, this cohort incorporates a broad spectrum of rare disorders recruited to the 100kGP study, with minimal pre-selection based on probable diagnostic candidates or phenotypic observations. Utilising the DROP framework, we identified gene expression and splicing outliers via OUTRIDER and FRASER2 respectively and applied multiple strategies to prioritise candidate diagnoses.

Several individuals were event count outliers, with high numbers of expression and/or splicing outliers. These individuals were found to be different for several related QC metrics, with the number of mapped unique reads and the number of genes detected in the data being the most significantly reduced in the event count outlier individuals relative to the rest of the cohort. This demonstrates the importance of high quality, high sequencing depth RNA-Seq for the reliable detection of both splicing and expression outliers. A greater sequencing depth is required for reliable outlier identification relative to group based differential expression or differential splicing analyses, which must be borne in mind during study design.

There was limited overlap in the genes identified as splicing outliers by FRASER2 and as expression outliers by OUTRIDER. This is despite many splicing events being expected to result in nonsense mediated decay of transcripts, which would be expected to lead to lower expression of the gene. This may partly be due to the sequencing methodology. Ribo-depleted libraries contain more immature transcripts than PolyA captured libraries (Zhao et al, 2018), so transcripts which are destined for nonsense mediated decay may be being captured before maturity. This underlies the importance of utilising both approaches to capture the full range of impacts at the RNA-level.

In several instances (e.g. *PHIP, FBX011*), RNA-seq provided additional evidence for variants previously classified as VUSs in the Genomics England diagnostic pipeline. VUS are a major bottleneck in clinical genomics, with more being identified as the scale of sequencing increases (Chen et al, 2023), creating uncertainty for clinicians and patients (Burke et al, 2022), and exacerbating existing inequalities in healthcare (Chen et al, 2023). RNA-Seq provides a relatively high throughput and efficient approach to resolving such variants (Dekker et al, 2023; Luo et al, 2025).

The Case Studies presented here represent a variety of different disease mechanisms and underlying variant types, illustrating RNA-Seq’s broad utility in rare disease diagnostics. For instance, a short 1.55kb deletion in *CTNNB1* which had been filtered out of the structural variant call set due to low quality, likely due to its small size, was detected as an exon skipping event by FRASER2. This highlights RNA-Seq’s ability to flag small deletions and CNVs affecting coding regions that are challenging for DNA sequencing technologies (Lincoln et al, 2021).

Several causal variants identified lay further from the intron:exon boundary than is typically included in variant prioritisation pipelines (Wright et al, 2015)(e.g. *WDR26*, *KMT2D, SPAST, DNMT3A*), which often target only canonical splice acceptor and splice donor sites, or may include “splice_region_variants” (spanning the first/last 3 bases of the exon, and up to 8bp of the intron). Historically, systematic assessment of intronic variants was limited by the lack of accurate, scalable and user friendly *in silico* tools for splicing prediction. The lower levels of conservation observed in introns mean there are relatively more variants in intronic regions, and a smaller proportion of those variants are expected to be causal. However, tools such as SpliceAI can now be integrated into variant prioritisation pipelines at scale, which would ensure more variants of this nature are captured and assessed clinically. All example variants were predicted to be splice disrupting by SpliceAI at a 0.2 cutoff, although more strict thresholds would have missed variants (e.g. *KMT2D* variant maximum SpliceAI=0.27).

In several instances, the gene harbouring the likely causal variants identified in this study had previously been suspected clinically. For example, the individual highlighted in Case Study 4 was recruited to the study under the classification of Kabuki syndrome. This is a highly clinically recognisable phenotype with a finite number of associated genes, predominantly *KMT2D.* The participant remained genetically undiagnosed despite strong clinical suspicion and GS, as the systematic assessment of intronic variants is not routinely undertaken. With a suggestive SpliceAI score of 0.27, this variant could have been identified sooner if reanalysis of the gene with *in silico* tools had been undertaken. In Case Study 7, there was a strong family history suggestive of *APC* variant involvement, and indeed, several family members had previously undergone genetic testing of the *APC* gene. Although not unique (Baumann et al, 2025; Miki et al, 1992), the unusual nature of the intronic retrotransposition event, and the complexities of resolving the event even with RNA-Seq, GS and long-read data make it unsurprising it had not been identified in previous analyses. This case highlights the diversity of disease mechanisms that an RNA-Seq led approach can identify.

Our results highlight the variable nature of the impacts that disruptive variants can have on measured expression levels. For two examples, OUTRIDER detected apparent overexpression of the affected gene (*FBX011*, FC=1.32 and *APC*, FC=1.58). These results appear counterintuitive when considering the loss of function mechanism suspected in both cases but are likely due to intron retention causing more reads to be attributed to the gene. We also observed lower than expected depletion of measured expression levels, best exemplified by the compound heterozygous variants in *INPPL1* (Case Study 9). Both the splice-impacting synonymous variant and the frameshift 1bp deletion would be expected to result in a frameshift effect, leading to nonsense mediated decay of the affected transcripts, yet the FC reported by OUTRIDER was just 0.72. These findings highlight that while it may be intuitively expected that a loss of function variant would result in a 50% depletion of the transcript being affected, the reality is much more complex and careful consideration of all findings, over and under expressed, is needed to capture all disease relevant findings.

The *SPAST* pseudoexon creation discussed in Case Study 5 was detected as an expression outlier (FC=0.74) rather than by splicing outlier analysis, despite the major impact of the variant being on splicing. The event had not reached genome-wide significance with FRASER2, likely due to the stop codon within the created pseudoexon triggering nonsense mediated decay, limiting the quantity of transcripts containing the pseudoexon sampled, and thus limiting statistical power. The *CTNNB1* frameshift-causing single exon deletion in Case Study 1 may be expected to impact expression, yet was not a significant outlier for expression and was identified through it’s apparent impact on splicing. This highlights the importance of utilising both splicing and expression analyses to capture the full range of disease relevant findings, even when prior expectations may suggest a variant would lead to a particular type of disruption.

Several factors limit the diagnostic yield of blood-based RNA-seq. Firstly, disease-relevant genes may be poorly expressed or absent in blood. Although approximately 60% of all PanelApp genes were well captured in the data, that leaves ∼40% of PanelApp disease associated genes poorly expressed at a TPM ≤ 5 in blood. Transcripts present in blood may also not be representative of the disease relevant tissues. Indeed, several studies have reported that relative to RNA derived from other clinically accessible tissues such as fibroblasts or muscle biopsies, blood derived RNA has lower and less consistent gene expression across disease relevant genes (Dekker et al, 2023; Gonorazky et al, 2019; Murdock et al, 2021; Rowlands et al, 2021; Yepez et al, 2022). However, blood, along with other tissues such as hair follicles and urine (Martorella et al, 2023), are more easily accessible so sampling may be more readily accepted by patients, as well as being simpler, faster, and cheaper to process. Several studies have demonstrated that blood-based RNA-Seq can be an effective way to identify diagnoses in patients with rare Mendelian disorders (Bertoli-Avella et al, 2025; Frésard et al, 2019; Jaramillo Oquendo et al, 2024). There was a high degree of variability in the proportion of relevant genes well captured by this dataset across different types of disorder. Over 90% of genes related to congenital disorders of glycosylation were captured at TPM ≥ 5, while for monogenic hearing loss and congenital myopathies, these values were considerably lower (35% and 32% respectively) In this paper we have provided a breakdown of the proportions of genes well captured in blood across different disease gene panels, which may be useful in guiding tissue of choice for future sequencing studies.

Secondly, our study mainly focussed on known, dominant, haploinsufficient genes which had been included in gene panels applied in the analysis for each participant. Recessive genes, non-haploinsufficient genes and genes which were not included in a participant’s relevant panels likely harbour additional diagnoses. We additionally did not explore novel disease gene discovery, which this resource offers exceptional promise for. Indeed, the resource has already been utilised in the discovery of two snRNA (*RNU4-2* and *RNU2-2*) related neurodevelopmental disorders (Chen et al, 2024; Jackson et al, 2025). Although non-coding genes are less well captured in the dataset than coding genes, it was used to demonstrate decreased *RNU2-2* expression in biallelic variant carriers (Jackson et al, 2025), and give insights into the mechanisms of disruption in each instance, with an increase in the use of unannotated splice donor sites in *RNU4-2* variant carriers (Chen et al, 2024), and an increase in shared aberrant splicing events in individuals with dominant *RNU2-2* variants (Jackson et al, 2025).

Third, the identification of gene expression and splicing outliers relied predominantly on two tools, OUTRIDER and FRASER2. These were selected as they are relatively widely used in the literature (De Cock et al, 2025; Dekker et al, 2023; Jaramillo Oquendo et al, 2024), and their incorporation into the DROP pipeline (Yepez et al, 2022) gives a scalable, relatively user-friendly approach for outlier identification. The use of additional tools and approaches to identifying outliers would also likely increase diagnostic yields. Particularly for splicing, different tools have been shown to identify different splicing events (Mehmood et al, 2019) and have different sensitivities and specificities for finding causal variants (Oquendo et al, 2024). There is currently no gold-standard for identifying gene expression and splicing outliers in the rare disease context, with a variety of different tools and approaches used in the literature (e.g. (Dekker et al, 2023; Frésard et al, 2019; Jaramillo Oquendo et al, 2024; Kremer et al, 2017)).

Comprehensive comparison of alternative methods to develop a reliable, user-friendly pipeline is paramount for the widespread adoption of RNA-Seq for rare disease diagnostics. Additional complexity stems from the impacts of splice-affecting variants often being incomplete or “leaky”, and there is currently no consensus on what level of disruption is sufficient to be considered pathogenic. Evidence that “leaky” splicing defects can lead to atypical disease presentations (e.g. (Ait-El-Mkadem Saadi et al, 2022; Li et al, 2019)) further complicates interpretation.

Finally, even for the outliers identified by the tools used, not all events could be comprehensively assessed for disease relevance. Manual evaluation is labour-intensive, requiring identification of associated variants, verification of effects on expression or splicing, prediction of protein-level consequences, and comparison of the gene’s known pathogenic phenotypes with the individual’s presentation. While we prioritised a set of the most likely causal events, additional relevant findings likely remain unidentified. Further research is needed to optimise methods for identifying, triaging and assessing the technical and clinical validity of outliers to expedite clinical translation.

We view this work as a starting point. This paired genome-transcriptome resource, now expanded to encompass over 7800 individuals with rare disorders, is freely available within Genomics England’s NGRL to bona fide registered academic researchers to increase diagnostic yields and bring new insights into rare disease aetiology. We hope a wide range of researchers with expertise in different disease types and genes will take advantage of this resource and contribute to this diagnostic uplift. It will also be a valuable resource for functional genomics research including into the regulation of gene expression and splicing, investigation of the effects of different variant types, and evaluation of the utility of blood-based RNA-Seq as a diagnostic tool. For many of these individuals, proteomics data, metabolomics and long read GS data is also available or being generated, providing a unique, rich multi-omics dataset which will catalyse improvements in diagnostics in rare disease as well as other applications.

In summary, we present a powerful dataset of RNA-Seq from over 5,000 individuals with rare disorders recruited to the 100kGP. The splicing and gene expression outlier datasets are available for use for researchers with access to the NGRL (see data access section below). We demonstrate the utility of this dataset in identifying new diagnoses, previously not identified for a wide variety of different reasons. We have identified significant splicing and/or gene expression outliers in potentially relevant disease genes in 20% of the cohort, representing many new potential diagnoses, and demonstrating the potential for further diagnostic uplift as this dataset is comprehensively explored over the coming years.

## Data Availability

Data from the National Genomic Research Library (NGRL) used in this research are available within the secure Genomics England Research Environment. Access to NGRL data is restricted to adhere to consent requirements and protect participant privacy. Data used in this research include:
RNA-Seq data (bam files, summary TPM data), GS data (bam files, VCF files), participant information (e.g. demographics, phenotype classes and HPO terms accessed through LabKey). Derived datasets produced include a table of 5412 participants included in the final analysis set with demographic and phenotype information, summary table of TPM across the cohort for all genes (including mean, median, min, and max values), summary tables of outlier splicing and expression events produced from DROP. These derived files are available to other researchers in the NGRL in /shared_allGeCIPs/RNA-Seq_outliers.
Access to NGRL data is provided to approved researchers who are members of the Genomics England Research Network, subject to institutional access agreements and research project approval under participant-led governance. For more information on data access, visit: https://www.genomicsengland.co.uk/research

https://doi.org/10.6084/m9.figshare.4530893

## Acknowledgements

We gratefully acknowledge the participants of the National Genomic Research Library (NGRL), whose contributions made this research possible. Secure access to the NGRL under project ID RR166 was provided by Genomics England, which delivers the NGRL in partnership with NHS England, and is wholly owned by the UK Department of Health and Social Care. The NGRL contains participants’ health data collected by the NHS as part of their care, along with samples and data from their participation in research, for which fully informed consent has been obtained. This includes genomic and clinical data provided through the NHS Genomic Medicine Service, as well as data obtained through research studies, including the 100,000 Genomes Project and the Generation Study, both of which are delivered in partnership with the NHS, and from other research cohorts involving external collaborators.

Cassandra Smith, bioinformatician in diagnostic discovery at Genomics England provided scripts to summarise the PanelApp gene lists which were used in the project. Dagmara Furmanczyk, Fabrice Kamole and Helena West (Illumina Cambridge Ltd, Granta Park) all provided vital support in sample processing and method validation in producing the RNA-Seq data at Illumina. Joseph M. Paggi provided support in the interpretation of the LaBranchoR scores for the *KMT2D* variant. Ian Eperon provided guidance and advice on early stages of the study and project design.

## Web Resources

Clinical Genome Resource, https://clinicalgenome.org

Genomics England 100,000 Genomes Project Exit Questionnaires, https://re-docs.genomicsengland.co.uk/exit_questionnaire/

SpliceAI score downloads, https://basespace.illumina.com/s/otSPW8hnhaZR

LabKey, https://re-docs.genomicsengland.co.uk/labkey/

The National Genomic Research Library, Genomics England (2024). https://doi.org/10.6084/m9.figshare.4530893

## Data access

Data from the National Genomic Research Library (NGRL) used in this research are available within the secure Genomics England Research Environment. Access to NGRL data is restricted to adhere to consent requirements and protect participant privacy. Data used in this research include: RNA-Seq data (bam files, summary TPM data), GS data (bam files, VCF files), participant information (e.g. demographics, phenotype classes and HPO terms accessed through LabKey). Derived datasets produced include a table of 5412 participants included in the final analysis set with demographic and phenotype information, summary table of TPM across the cohort for all genes (including mean, median, min, and max values), summary tables of outlier splicing and expression events produced from DROP. These derived files are available to other researchers in the NGRL in /shared_allGeCIPs/RNA-Seq_outliers.

Access to NGRL data is provided to approved researchers who are members of the Genomics England Research Network, subject to institutional access agreements and research project approval under participant-led governance. For more information on data access, visit: https://www.genomicsengland.co.uk/research

