## Supplemental Information for "Blood-based RNA-Seq of 5412 individuals with rare disease identifies new candidate diagnoses in the National Genomic Research Library"

1. **Supplemental Case Studies**
2. **Supplemental Notes**
3. **Supplemental Tables and Figures**
4. **Supplemental References**

**1. Supplemental Case Studies**

**FRASER2 splicing outliers in ClinGen Haploinsufficient Genes**

**Case Study 1**: *CTNNB1* splicing outlier flags undetected single exon deletion

A splicing outlier event in *CTNNB1* was detected in a male proband recruited under the Intellectual Disability disease group. *CTNNB1* encodes β-catenin, a protein with important roles in cell adhesion and cell signalling. Variants in *CTNNB1* are associated with neurodevelopmental disorder with spastic diplegia and visual defects (MIM:615075), a syndromic condition characterised by global developmental delay, intellectual disability, microcephaly and commonly with visual abnormalities that can include strabismus, optic nerve atrophy, hypermetropia and exudative vitreoretinopathy (de Ligt et al, 2012; Tucci et al, 2014). The phenotype in the proband with the *CTNNB1* outlier included intellectual disability, developmental delay, spasticity and microcephaly, and was thought to be consistent with the phenotype associated with loss of function variants in the gene.

The splicing abnormality detected was annotated by FRASER2 as an exon skipping event (deltaPSI=-0.32, gene level adjusted p-value=3.82x10^-5^). Skipping of exon 14 (ENST00000349496) was clearly apparent upon manual inspection using IGV, with 65 reads supporting the skipping of the exon (**Figure 3ai**). Exon 13-14 splicing was supported by 105 reads, and exon 14-15 splicing was supported by 83, giving an aberrant splicing ratio of approx. 0.69 (65/((105+83)/2)). Exon 14 is 61bp, so skipping is expected to cause a frameshift effect and loss-of-function of the allele. Although *CTNNB1* was not detected as a significant gene expression outlier for this individual after multiple test correction, modest reduction in transcription was observed (fold change (FC)=0.88, uncorrected p-value of 0.00037, OUTRIDER).

Inspection of the RNA-Seq sequencing reads, and interrogation of the individual’s GS VCFs containing SNVs, short indels and high quality CNVs did not reveal any variants which could explain the observed splicing event. However, manual inspection of the individuals’ DNA sequencing reads in IGV revealed a short, 1.55kb deletion spanning the “skipped” exon 14, with evidence of CT homology at each breakpoint (chr3:41236871-41238422 (GRCh38), **Figure 3aii**). This event was called by Manta but had been filtered as a low quality CNV call (q10, CLT10kb), likely due to its small size, but is clearly apparent from the DNA sequencing read data.

**Case Study 2**: RNA-Seq clarifies impact of *PHIP* variant of unknown significance

A splicing outlier event was identified in the gene *PHIP* (deltaPSI=-0.48, adjusted p-value=0.009). Manual inspection of the region in IGV showed skipping of exon 6 (ENST00000275034), with a donor+2 splicing variant apparent at chr6:79060476:A:C (GRCh38, **Figure 3b**). This ENST00000275034.5:c.439+2T>G variant had been previously identified by the Genomics England diagnostic pipeline and had been classified as a variant of unknown significance (VUS). The clear evidence of exon skipping provided by the RNA-Seq demonstrates the impact of the variant at the RNA level and this is predicted to result in an in-frame removal of 34 amino acids and insertion of a threonine residue, p.(Ser114_Ala147delinsThr). SpliceAI (Jaganathan et al, 2019) scores (**Figure S6a**) predict the variant would cause exon skipping, with loss of the splice donor (0.98) and acceptor site (0.93). Pathogenicity is supported by a nearby *de novo* variant (c.439+5G>T) affecting the same splice donor site listed in ClinVar as likely pathogenic (VCV002664249.1) and for which SpliceAI also predicts skipping of exon 6.

Pathogenic variants in *PHIP* are associated with the autosomal dominant Chung-Jansen syndrome (MIM:617991), characterised by developmental delay, intellectual disability, obesity and dysmorphism (Webster et al, 2016). This is consistent with the HPO terms recorded for our proband, which include intellectual disability, developmental delay and prominent forehead.

**Case Study 3**: Intronic variant 50bp from *WDR26* splice site causes cryptic site activation

A splicing outlier event (exon elongation, deltaPSI=0.18, adjusted p-value=0.02) was identified in *WDR26* in an individual with intellectual disability, developmental delay and seizures. As this presentation would be consistent with Skraban-Deardorff syndrome (MIM:617616), manual inspection of the region in IGV was performed. This showed the use of a cryptic splice donor site within the intron near exon 8, leading to the inclusion of 51bp of intronic sequence in transcripts (**Figure 3c**). Although this is an in-frame exon elongation event, a premature termination codon is encoded in the additional sequence. Use of this alternative event is supported by 37 sequencing reads, versus 92 which show the use of the annotated exon 8 splice donor site.

The *de novo* ENST00000414423.9:c.1599+51T>G variant which activates the new splice site (chr1:224404379:A:C (GRCh38)) falls in the position preceding an existing intronic “GT” dinucleotide, which forms the new cryptic splice donor. The nucleotide before the splice site “GT” dinucleotide is most often a G, and has been shown to be resistant to variation (Lord et al, 2019). The final base of exon 8 (the equivalent position in the annotated splice site) is an A, giving a weaker match to the splicing consensus sequence than this newly created intronic splice site. Indeed, MaxEntScan (Yeo & Burge, 2004) scores the real exon 8 splice donor site as weaker than the new intronic cryptic splice site (6.39 vs 7.87 respectively), and SpliceAI scores (**Figure S6b**) support the activation of a new splice donor site (donor gain=0.83) and weakening of the annotated exon 8 donor site (donor loss=0.50).

Although this variant is a *de novo* variant in a known disease gene relevant to the proband’s phenotype, its location >50bp from the splice donor site means it was overlooked as a potential causal candidate through GS analysis alone, since this typically only includes up to 8bp of intronic sequence as the “splice region”.

**Case Study 4**: *KMT2D* intronic variant likely disrupts branchpoint

A partial intron retention event was detected in *KMT2D* (deltaPSI=-0.37, adjusted p-value=1.22x10^-9^) in an individual recruited with suspected Kabuki syndrome. Consistent with the phenotype observed in individuals with pathogenic variants in *KMT2D* (MIM:147920), the case’s phenotype included abnormalities of the genitourinary system, the head or neck, eye, ear and limbs, as well as intellectual disability and global developmental delay. Manual inspection of the region in IGV confirmed complex splicing disruptions, including intron retention and the use of multiple cryptic splice sites (**Figure 3d**).

Two potential variants were visible in the RNA-Seq reads at chr12:49042333:G:A and at chr12:49042352:T:G, but only the latter of these was present in the individual’s DNA VCF, and evidence of the alternative allele at the chr12:49042333 position could be observed in all control data checked, suggesting this is a RNA-Seq artefact and not a genuine variant. The proband was recruited as part of a duo, with the variant not being present in the maternal DNA sequencing data, and with no paternal data available.

The proband’s candidate pathogenic variant, chr12:49042352:T:G (ENST00000301067.12: c.5868-22A>C), is 22bp upstream of the splice acceptor site of exon 29. Although SpliceAI only predicts a modest impact on the strength of the splice acceptor site (acceptor loss=0.27, **Figure S6c**), the variant likely disrupts the critical A nucleotide of the branchpoint for the splice site (the gene is on the negative strand). Indeed, LaBranchoR (Paggi & Bejerano, 2018) identifies this position as the branchpoint for this exon, and predicts strong disruption of the branchpoint (d_best_bp score=-0.55323).

This was not identified as a potential diagnosis in the primary analysis as it is annotated as an intronic variant which are typically filtered out in GS analysis.

**Diagnostic candidates identified from analysis of OUTRIDER expression outliers**

**Case Study 5**: *SPAST* pseudoexon creation in two families with hereditary spastic paraplegia

An additional splice-disrupting variant was identified through analysis of the OUTRIDER gene expression outliers, rather than FRASER2 (the region was flagged by FRASER2, but the p-values did not withstand correction for multiple testing). An individual recruited with hereditary spastic paraplegia (HSP) was found to have significantly reduced expression of *SPAST* (FC)=0.75, adjusted p-value=0.00026). Given this gene is linked to autosomal dominant spastic paraplegia type 4 (MIM:182601), we searched for rare variants including in intronic reqions and identified a heterozygous chr2:32128139:C>T (ENST00000315285.9:c.1174-269C>T) variant 268bp from the exon 9 splice acceptor site, for which SpliceAI predicts a 120bp pseudoexon (acceptor gain=0.62, donor gain=0.66, **Figure S6d, S7a**). Although absent from gnomAD v4.1.0, this variant was found in the 100kGP aggregate VCF file (AggV2) at an allele frequency of 5/156,390. The 5 heterozygotes were from two families, both recruited with HSP. Individuals with HSP make up just 0.68% (528/78,195) of the AggV2 cohort.

In the first family, the variant was present in both the proband and his affected sister. These individuals showed a slowly progressive adult-onset lower limb spasticity phenotype with bladder involvement but no upper limb involvement. In the second family, two sisters were recruited with HSP. The variant had been inherited from the father who had not been formally assessed. For both sisters, RNA-Seq was available, and data supported the introduction of a pseudoexon (**Figure S7b**), consistent with the *in silico* prediction. The pseudoexon contains a premature stop codon (NP_055761.2:p.(A392Tfs*14)) which would likely lead to nonsense mediated decay of transcripts containing the pseudoexon, reflected in the FC=0.75 identified by OUTRIDER. This would result in a loss of function impact well documented as a cause of HSP (MIM:182601).

**Case Study 6**: large deletion linked to *RPL5* gene expression outlier

In an individual recruited to the 100kGP under the category of congenital anaemias, OUTRIDER identified a gene expression outlier in the gene *RPL5* (FC=0.56, adjusted p-value=2.82x10^-12^). Disruption of *RPL5* causes Diamond-Blackfan anaemia 6 (MIM: 612561), a syndromic anaemia with onset typically in the first year of life (Gazda et al, 2008). Cross-referencing with SV calls revealed a large deletion spanning 927kb, which was identified by both Manta and Canvas. Manta reports the genomic coordinates to be chr1:91907874-92835348 (GRCh38), although this is flagged as imprecise, likely due to the presence of Alu elements at the respective breakpoints.

In addition to *RPL5*, this deletion included seven other protein coding genes, three of which were also significant OUTRIDER outliers (*GLMN,* FC=0.41, adjusted p-value=2.52x10^-5^; *RPAP2*, FC=0.56, adjusted p-value=2.44 x10^-16^; *EVI5,* FC=0.53, adjusted p-value=5.24x10^-7^). Of the remaining four genes, two were not assessed by OUTRIDER (*BRDT, EPHX4*), presumably owing to low coverage (median TPMs across the cohort of 0.0 and 0.28 respectively), while two were assessed by OUTRIDER but were not significant (*BTBD8,* FC=0.64, adjusted p-value=1, TPM=2.99; *GFI1*, FC=0.95, adjusted p-value=1, TPM=12.17), which is thought to be due to low expression (*BTBD8*) or the relatively small size of gene (*GFI1,* 13kb) limiting power (**Figure S9**).

**Case Study 7:** intronic retrotransposon in *APC*

In an individual recruited to the 100kGP due to multiple bowel polyps. There was significant family history, with the index proband being one of 8 affected individuals spread across three generation of a single kindred. Prior to the proband’s recruitment, three other individuals had undergone testing for variants in *APC* between 2008 and 2014, with no diagnostic findings being uncovered.

RNA-Seq data showed significantly upregulated expression of *APC* (FC=1.58, adjusted p-value=6.91x10^-25^), with much of this signal localised to intron 10. Again, although FRASER2 did not detect any splicing abnormalities at genome-wide significance, an event was flagged at this affected intron, but this did not survive correction for multiple testing. Although no structural variants were called by Manta at this locus, scrutiny of this intron using short-read GS and RNA-Seq data indicated a potential breakpoint, where split-read pairs were identified, with one of the split-reads mapping to a centromeric repeat region on chromosome 17 (**Figure S10**). As several reads supported a polyA insertion and the integration site appeared to be at a TTAAAA motif (which is known to be the preferred cut site of the L1 endonuclease), we speculated that a retrotransposition event had taken place.

Using nanopore long-read DNA sequencing, we obtained 10 reads which spanned both ends of the insertion and the 1.9kb sequence matched that of an SVA type D retrotransposon. Using a common heterozygous SNV in exon 14 (rs351771; p.Ala545=) we were able to separate the two haplotypes (**Figure S11a**) and using long-read RNA-Seq showed that aberrant splicing was exclusive to the haplotype harbouring the SVA integration (**Figure S11b**). We note that SVA insertions in intron 9 of *APC* were described recently (Nakamura et al, 2024) but where nanopore sequencing was used for initial detection rather than by the RNA-first approach used here. Validation of the SVA insertion was performed using a PCR-Sanger sequencing approach (**Note S1, Figure S12**) and cascade testing employed a 3-primer approach, which showed that 4/4 of the proband’s available affected relatives also harboured the insertion (**Figure S13**).

**Exomiser identified diagnostic candidates**

**Case Study 8**: *FBXO11* missense variant impacts splicing and expression

A *de novo* missense variant in *FBXO11* (chr2:47813234:C:T, (GRCh38); ENST00000403359.7:c.2227G>A:p.Gly743Ser) was identified in a proband recruited under the Intellectual Disability disease group. The individual presented with global developmental delay, intellectual disability, delayed speech and language development, autistic behaviour, fine and gross motor delay, microcephaly, abnormality of the mouth, and digital anomalies such as 5th finger clinodactyly.

Exomiser ranked the *FBXO11* variant as the top candidate with a combined score of 0.948. This included a phenotype score of 0.726, driven by the strong similarity between the proband’s clinical features and those associated with *FBXO11*-related autosomal dominant intellectual developmental disorder with dysmorphic facies and behavioural abnormalities (MIM 618089), a condition in which numerous pathogenic *de novo* missense variants have been described. The variant also had a high variant score of 0.940 and was predicted to be deleterious by multiple *in silico* tools (REVEL (Ioannidis et al, 2016): 0.878; SpliceAI: 0.950, **Figure S6e**). The variant falls in the highly conserved last nucleotide of exon 18 (NM_001190274.2), just adjacent to the canonical donor site, and is absent from gnomAD v4.1.0.

OUTRIDER identified *FBXO11* as a gene expression outlier, with a relative increase in gene expression (FC=1.32, adjusted p-value=3.21x10^-13^). Analysis with SpliceAI and Pangolin (Zeng & Li, 2022) predicted the variant would result in skipping of the exon (**Figure S6e**). This impact was observed in the Sashimi plot (**Figure 4a**) and was detected by FRASER2 (deltaPSI=-0.45, unadjusted p-value=2.45x10^-6^) but did not remain significant after correction for multiple testing (adjusted p-value=0.179). We also observed an increase in intron retention apparent in the data, which is likely to explain the apparent increase in expression of the gene, despite disruption to splicing which is often associated with decreased expression.

This variant had already been assessed through the Genomics England diagnostic pipeline and was classified as a VUS. However, the additional evidence from RNA-Seq expression and splicing analysis supports a functional impact and may warrant reclassification of this variant as likely pathogenic.

**Case Study 9:** Compound heterozygous variants in *INPPL1*

Compound heterozygous variants in *INPPL1* were identified in an individual recruited under the chondrodysplasia punctata disease group. The individual presented with short stature and skeletal abnormalities. The variants were inherited in trans, from unaffected parents. Exomiser prioritised *INPPL1* as the top-ranked gene (Exomiser score: 0.830), with a phenotype score of 0.636, reflecting the overlap between the proband's features and the clinical features consistent with *INPPL1*-realted opsismodysplasia (MIM:258480), and a variant score of 0.900.

The first variant is a synonymous splice region variant (chr11:72234615:G:A, GRCh38; HGVS: ENST00000298229.7:c.2415G>A:p.=), which is located in the conserved last nucleotide of exon 21 (NM_001567.4). The variant is rare in GnomAD v4.1.0 (allele frequency: 0.000004352) and has a SpliceAI score of 0.52, predictive of a splicing impact on the adjacent canonical donor site (**Figure S6f**). A compound heterozygous variant at the adjacent nucleotide has been previously reported in individuals with opsismodysplasia (Below et al, 2013).

FRASER2 identified a significant deviation in splicing for *INPPL1* (deltaPSI=-0.61; adjusted p-value=2.85×10⁻⁶), with manual inspection in IGV confirming the splicing impact (**Figure 4b**). None of the variant-containing reads used the canonical splice site, with most utilising a cryptic splice donor site 31bp from the canonical acceptor site (**Figure 4bii**). This would lead to the inclusion of 31bp of intronic sequence in the transcript, leading to a frameshift effect.

The second variant is a frameshift variant (1bp deletion, chr11:72237532:GC:G, GRCh38 ; HGVS: ENST00000298229.7:c.3290del:p.Pro1097Leufs*34) in exon 26 (NM_001567.4). This variant is also rare (GnomAD frequency: 0.000006851) and within the same exon as five distinct pathogenic frameshift variants in ClinVar.

Expression analysis using OUTRIDER revealed *INPPL1* to be a gene expression outlier in the proband (FC=0.72, adjusted p-value=3.15×10⁻⁴). It is unclear whether this impact on expression arose from the splicing variant, the frameshift variant, or both, although it is noteworthy that the impact is modest despite two apparent loss of function variants which would be expected to include nonsense mediated decay.

Together, these findings support the conclusion that these biallelic variants disrupt normal *INPPL1* function. Although the splice variant was classified as a VUS after feedback from the GMC in the Genomics England pipeline, the gene’s known association with autosomal recessive opsismodysplasia and the strong phenotypic match, combined with transcriptomic and *in silico* evidence, support the reclassification of this variant from VUS to likely pathogenic.

**Case Study 10:** Homozygous *DYSF* missense variant alters splicing in individual with limb-girdle muscular dystrophy

A splicing outlier event in *DYSF* was identified in an individual presenting with axial muscle weakness, limb muscle weakness, and limb-girdle muscular dystrophy. These features are consistent with Muscular Dystrophy, Limb-Girdle, Autosomal Recessive 2 (LGMDR2, MIM:253601), caused by biallelic variants in *DYSF*.

The proband was homozygous for a missense variant in *DYSF* (chr2:71681110:G:A, GRCh38; HGVS: ENST00000410020.8:c.6173G>A:p.Arg2058Lys). The variant occurred in the last nucleotide of exon 54 (NM_001130987.2), adjacent to the canonical donor site. It is extremely rare in the general population, having been observed in only a single individual in gnomAD, corresponding to a frequency of 6.196x10⁻⁷. It was prioritized by Exomiser with a combined score of 0.637 (phenotype score: 0.771; variant score: 0.636). The variant was only ranked as 16^th^ as the proband was recruited as a singleton (therefore having a lot of variants that could not be filtered out by segregation) and the variant had relatively low pathogenicity scores (REVEL = 0.256; SpliceAI = 0.210, **Figure S6g**). The variant has conflicting interpretations of pathogenicity in ClinVar (one likely pathogenic, one VUS) although splice donor variants affecting the adjacent canonical donor site have been classified as likely pathogenic with multiple submissions.

Transcriptomic analysis using FRASER2 identified *DYSF* as a highly significant splicing outlier (deltaPSI=–0.92, adjusted p-value=1.30x10⁻⁴⁹). Manual inspection of RNA-Seq data showed the use of a cryptic splice donor site 143bp downstream of the canonical site, at position chr2:71681252 (NM_001130987.2), as well as exon skipping of the exon containing the variant (**Figure 4c**). SpliceAI and Pangolin predict moderate weakening of the existing splice donor (ΔSpliceAI = 0.21; ΔPangolin = –0.28) and acceptor sites (ΔSpliceAI = 0.16; ΔPangolin = -0.06), and strengthening of the cryptic site (ΔSpliceAI = 0.42; ΔPangolin = 0.16, **Figure S6g**). OUTRIDER identified *DYSF* as having moderate but non-significant downregulation (FC=0.7, adjusted p-value=0.9).

Taken together, the autosomal recessive mode of inheritance, strong phenotypic concordance with *DYSF*-related muscular dystrophy, and the clear splicing disruption demonstrated by transcriptomic analysis, these findings provide compelling evidence supporting the pathogenicity of this variant.

**Case Study 11:** *DNMT3A* intronic variant causes pseudoexon inclusion

After expanding the search area beyond 50bp from the intron:exon boundary, a splicing outlier event in *DNMT3A* was detected in an individual recruited under the Intellectual Disability disease group. They presented with global developmental delay, intellectual disability, macrocephaly, tall stature, autistic behaviour, and delays in speech, fine motor, and gross motor development, consistent with Tatton-Brown-Rahman syndrome (TBRS, OMIM: 615879), a condition associated with autosomal dominant *de novo* variants in *DNMT3A*.

The proband was found to carry a heterozygous variant in *DNMT3A* (chr2:25240802:T:C, GRCh38; HGVS: ENST00000321117.9:c.2083-72A>G:p.=), annotated as an intronic variant located 72 bp from the nearest exon–intron boundary. This variant was prioritised by Exomiser with a combined score of 0.823 (phenotype score: 0.640; variant score: 0.890), ranking *DNMT3A* as the second-best candidate gene for this individual. The variant is rare (gnomAD frequency 0.000009379) and is supported by a high SpliceAI score of 0.890, suggesting a splicing impact is likely (**Figure S6h**). Paternal data was not available, so although the variant was confirmed not to be inherited from the mother, confirmation of *de novo* inheritance was not possible.

FRASER2 identified *DNMT3A* as a significant splicing outlier (deltaPSI=–0.43; adjusted p-value=0.010778). Manual inspection of RNA-Seq reads using IGV revealed that the intronic *DNMT3A* variant creates a strong cryptic intronic donor site at the variant position (**Figure 4d**), leading to the creation of a 93bp pseudoexon between exons 17 and 18 (NM_022552.5).

Although this variant lies in a known dominant disease gene that is highly relevant to the proband’s phenotype, its position more than 70bp from the canonical splice donor site led to it being overlooked as a potential causal candidate. However, the novel transcriptomic evidence of splicing disruption, together with the strong phenotypic overlap, provides compelling support for the pathogenicity of the variant via a splice-altering mechanism.

**2. Supplemental Notes**

**Note S1: Validation of SVA insertion in *APC***

PCR reactions were performed using 19.3µl of MegaMix (Microzone), with 0.1µl of each primer per reaction (these were a 1 in 10 dilution of a 100µM stock). Also, 0.5µl of DNA at ~150ng/µl was used as template. Thermocycling conditions were 94°C for 5mins, then 35 cycles of 94°C for 30 seconds, 55°C for 1 min, 72°C for 1 min. A final extension was at 72°C for 5 mins.

The multiplex reaction used for cascade testing comprised primers 2025.427, 2025.438 and 2025.437. The upstream junction was determined using primers 2025.427/2025.428. The downstream junction was determined using primers 2025.437/2025.438.

**Primer set for control amplicon**

APC_dnstr_Rev2: AGCCAAAGAAACAATCCAGGA (2025.438)

APC_upstr_Fwd2: TCAGCCTATTGATGTGAAGAG (2025.427)

(product size: 363bp)

**Primer set for downstream junction**

APC_dnstr_Fwd2: GCTGACCTTCCCTCCACTAT (2025.437)

APC_dnstr_Rev2: AGCCAAAGAAACAATCCAGGA (2025.438)

Expected product size ~291bp

**Primer set for upstream junction:**

APC_upstr_Fwd2: TCAGCCTATTGATGTGAAGAG (2025.427)

APC_upstr_Rev1: CCTCCGAGACAGGGTTGC (2025.428)

Expected product size: ~403bp

**3. Supplemental Tables and Figures**

**Table S1:**  Separate Excel sheet (TableS1_ClinGenHI) listing 376 ClinGen Haploinsufficient genes.

**Figure S1:** Summary of QC information on the 5412 RNA-Seq samples in the final analysis set. A) Median number of mapped unique reads per individual. B) Median proportions of reads mapping to different genomic locations (NA = ambiguous). C) Median proportions of reads mapping to different GENCODE “gene_type”s, showing only those categories with >5 genes expressed at a TPM ≥ 5.

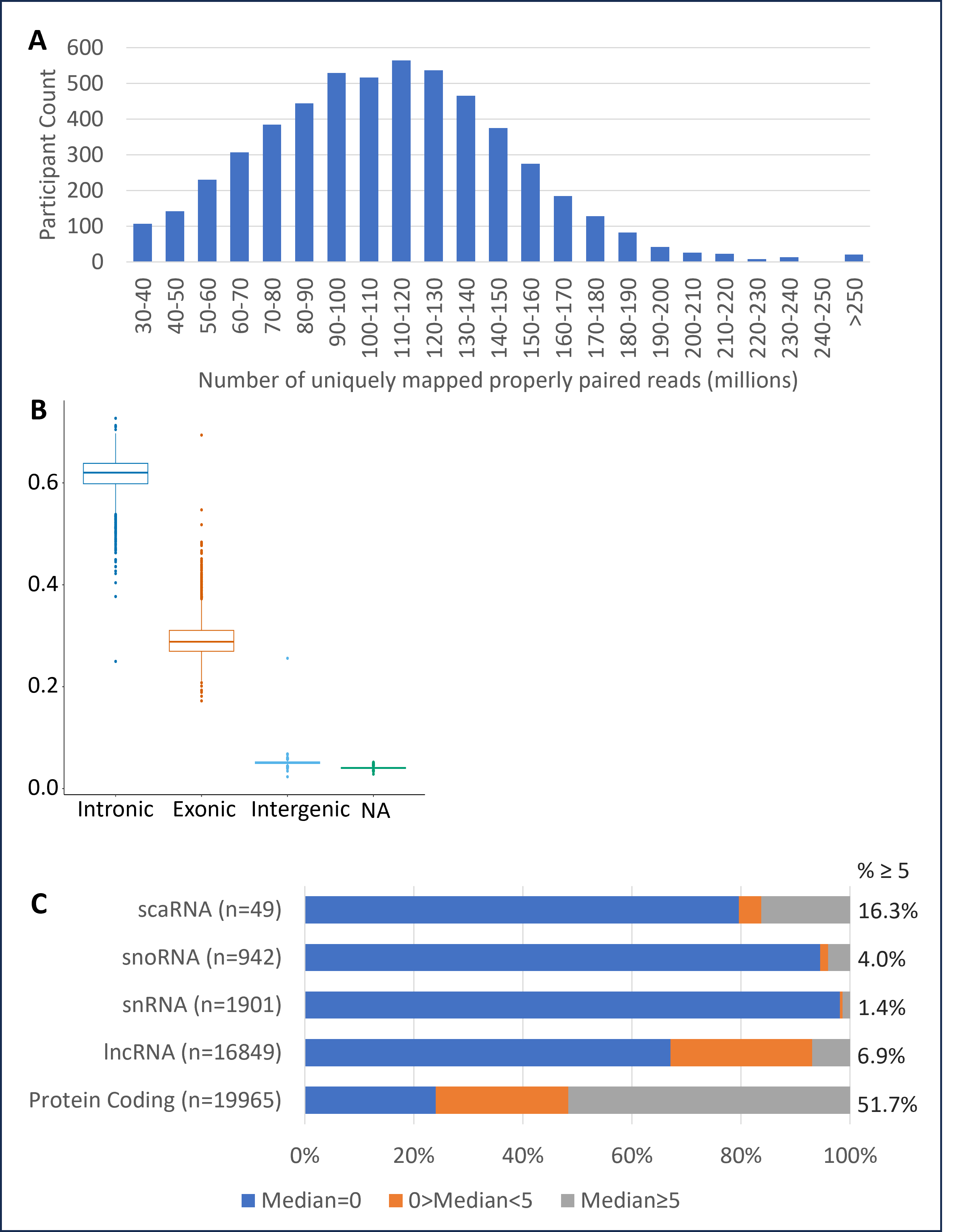

**Figure S2:** Number of outlier events identified by FRASER2 and OUTRIDER across the cohort across all genes.

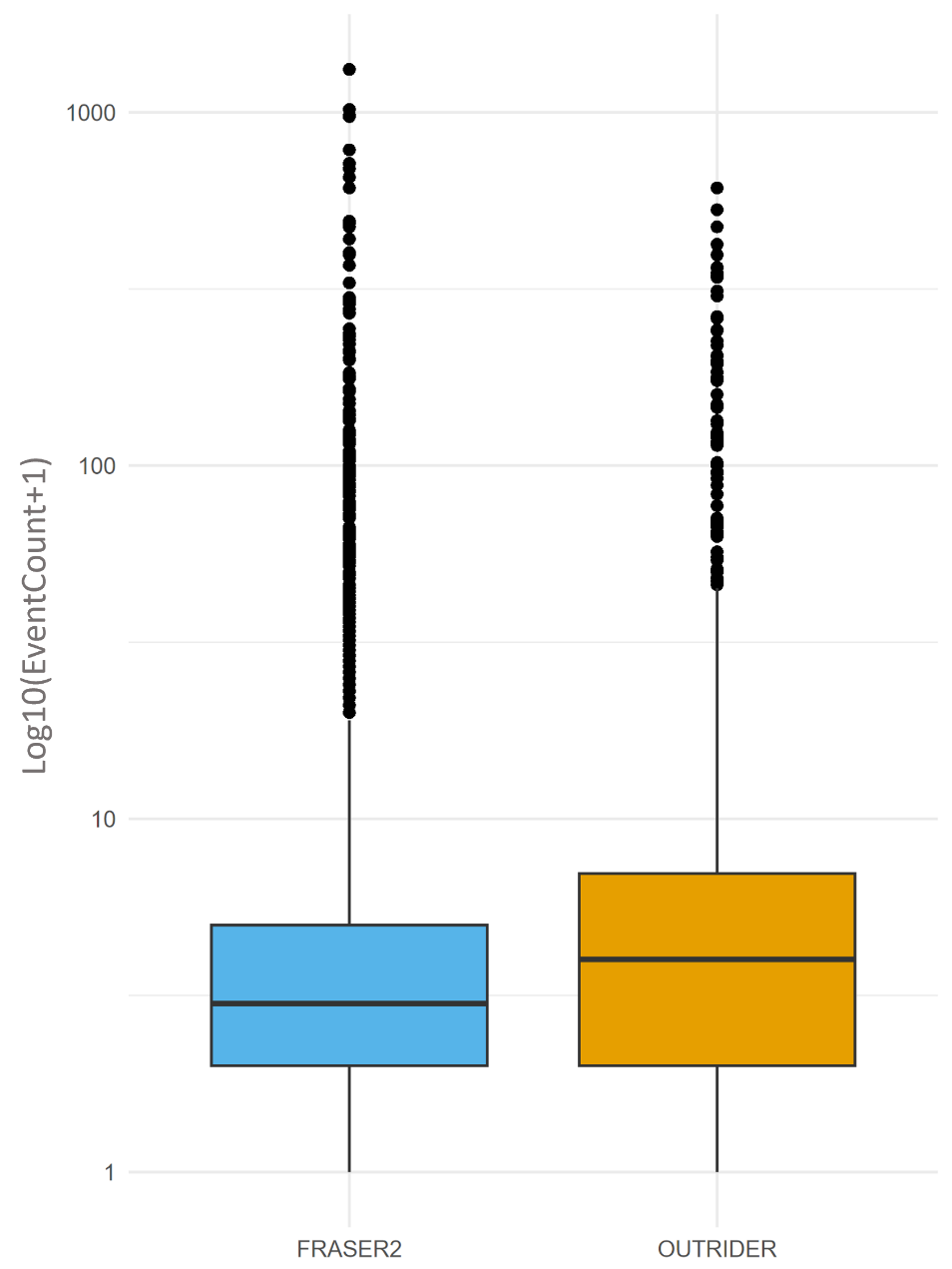

**Table S2:** Separate Excel sheet (TableS2_EventCountOutlier_stat) Wilcoxon test results for difference in RNA-SeQC2 derived metrics for FRASER2 and OUTRIDER event count outliers and the rest of the cohort, with FDR correction.

**Figure S3:** Boxplots showing difference in RNA-SeQC2 derived metrics for FRASER2 and OUTRIDER event between event count outliers and rest of the cohort. Five most significantly different metrics for each tool shown. cv = coefficient of variation, MAD = median absolute deviation.

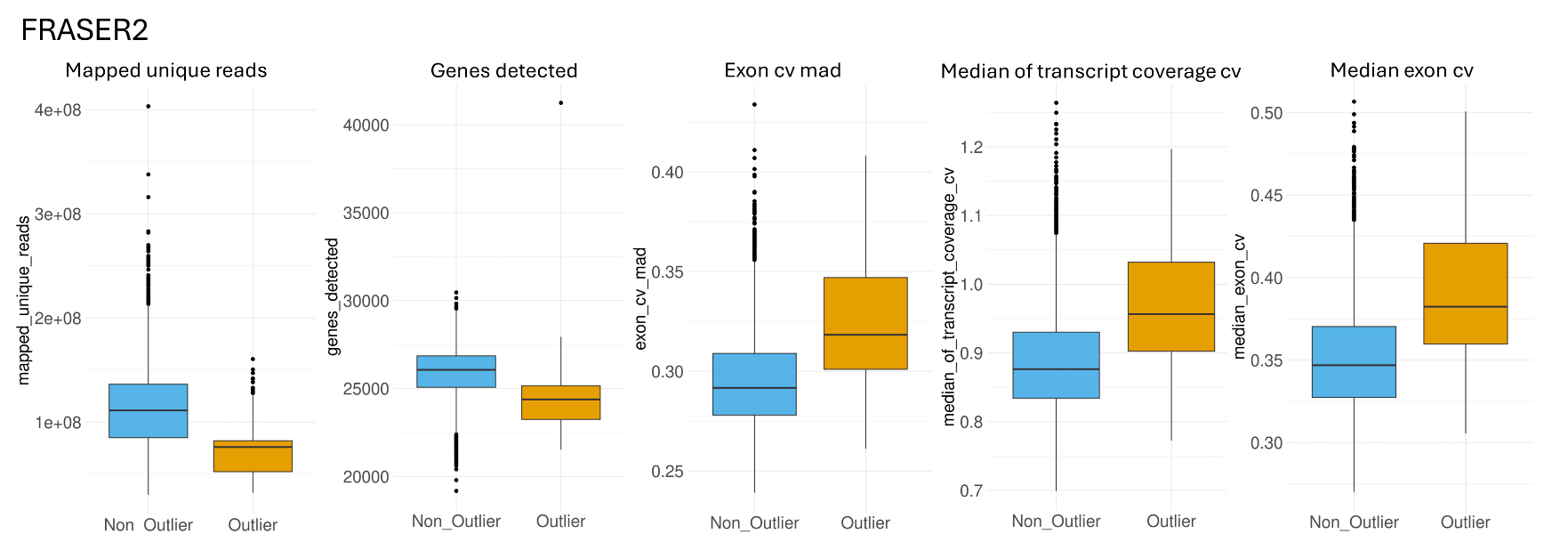

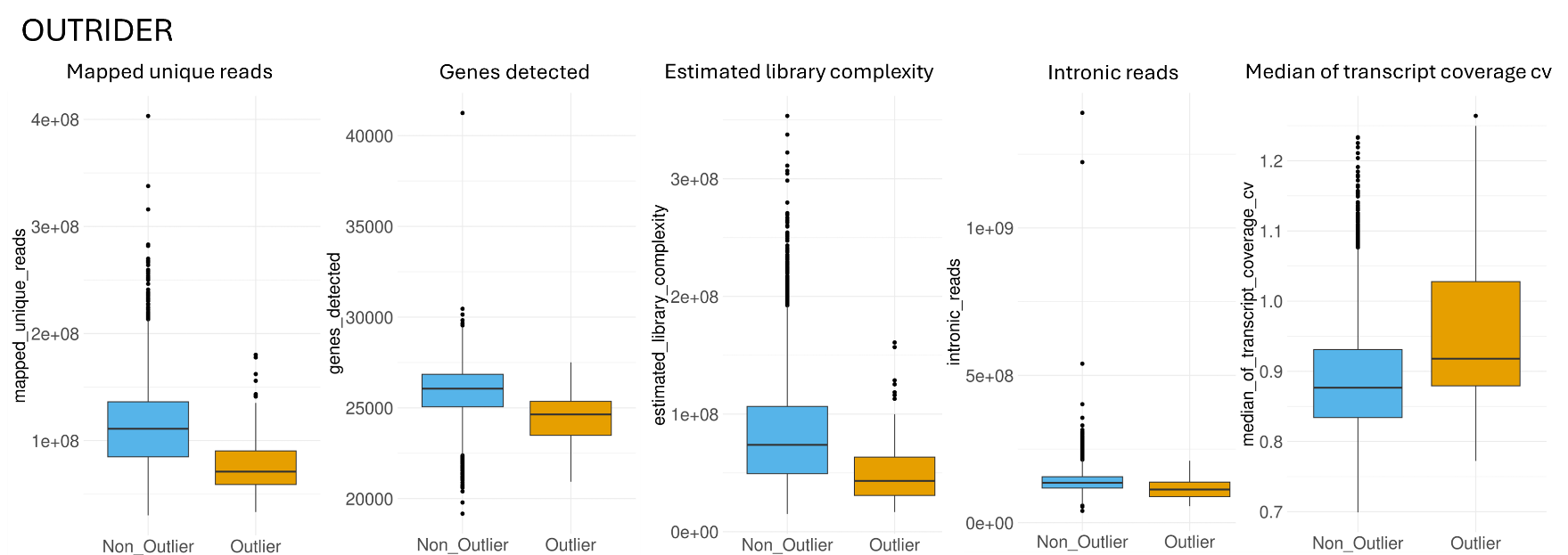

**Table S3:** Separate Excel tab (TableS3_PositiveControls) giving information on the 47 positive controls (individuals with genetic diagnoses from GS, where splicing impact was manually confirmed in the RNA-Seq data).

**Table S4:** Properties of the positive control variants that resulted in genome wide significant and nominally significant FRASER2 outlier events

| Metric | Significant | Nominal | p-value (t-test or *Fisher’s exact test) |
| --- | --- | --- | --- |
| Mapped Unique Reads (median) | 138.8888 | 113.734 | 0.1504 |
| Absolute DeltaPSI (median) | 0.48 | 0.31 | 0.000993 |
| Cohort gene TPM (median) | 126.0985 | 42.87725 | 0.009928 |
| Proportion with negative deltaPSI | 0.826087 | 0.875 | 0.7008* |
| Proportion OUTRIDER signficicant | 0.565 | 0.5 | 0.7725* |
| OUTRIDER raw p-value (median) | 0.004238 | 0.098617 | 0.6836 |
| OUTRIDER adjusted p-value (median) | 1 | 1 | 0.715 |
| OUTRIDER Fold change (median) | 0.99 | 0.94 | 0.8176 |
| OUTRIDER Proportion downregulated | 0.521739 | 0.708333 | 0.2375* |

**Figure S4:** Comparison of metrics between positive control events that resulted in genome wide significant and nominally significant FRASER2 outlier events

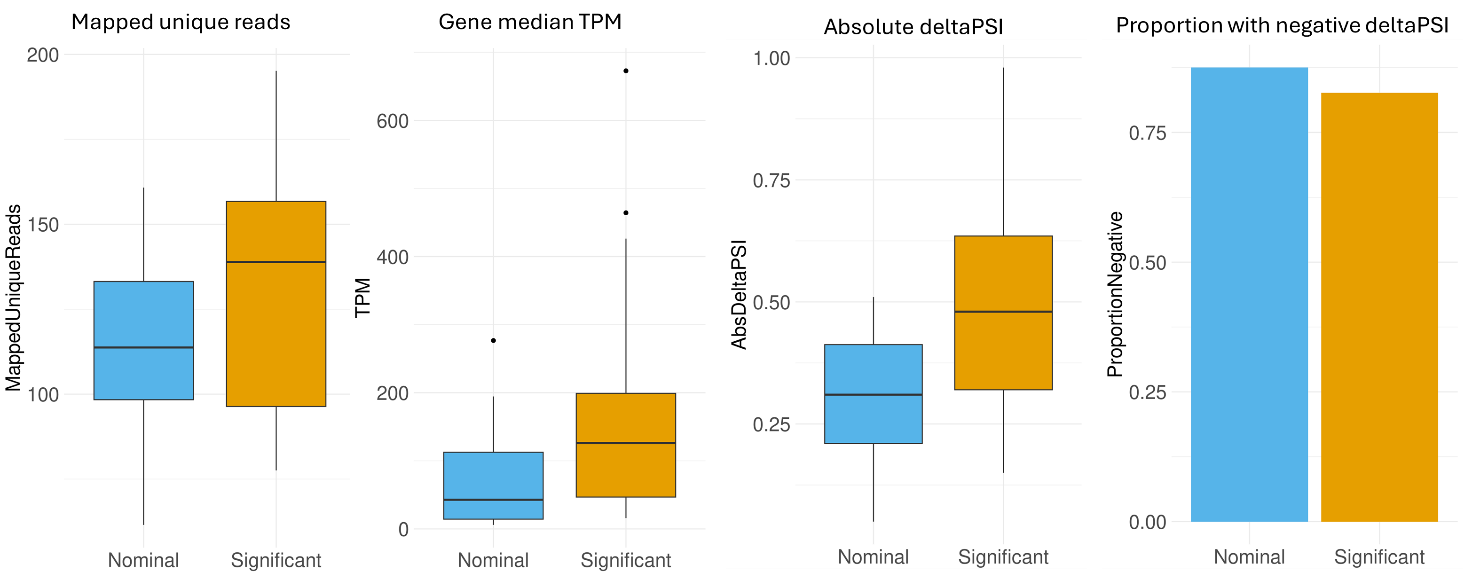

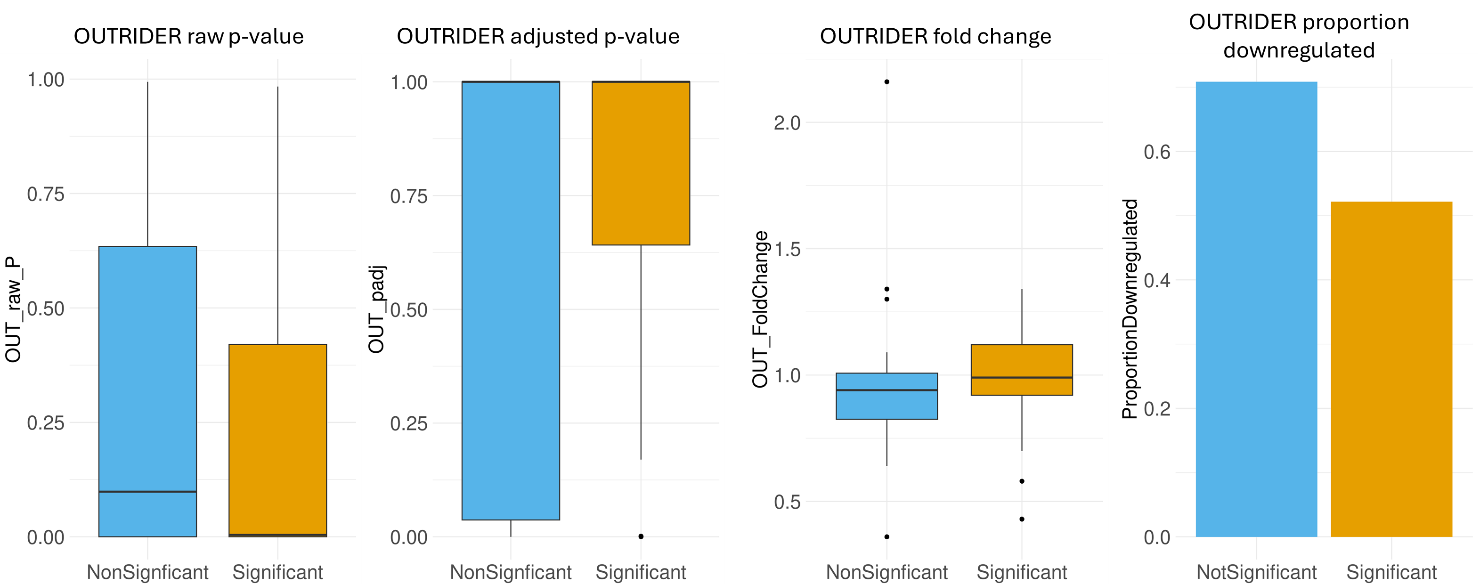

**Table S5:** Properties of FRASER2 splicing outlier candidates in ClinGenHI genes that passed or failed manual verification using IGV

|  | | **N events** | **Manually verified** | **Not verified** | **Unclear** |
| --- | --- | --- | --- | --- | --- |
| Events | 200 | | 78 | 70 | 52 |
| Median mapped unique reads | 200 | | 95.781 | 71.885 | 72.138 |
| Median gene level adjusted p-value | 200 | | 0.003 | 0.029 | 0.013 |
| Median absolute deltaPSI | 200 | | 0.4 | 0.335 | 0.395 |
| Proportion with negative deltaPSIs | 200 | | 0.641 | 0.486 | 0.712 |
| **FRASER2 event types:** |  | |  |  |  |
| annotatedIntron_reducedUsage | 62 | | 0.435 | 0.21 | 0.355 |
| exonSkipping | 11 | | 0.545 | 0 | 0.455 |
| (partial)intronRetention | 48 | | 0.396 | 0.333 | 0.271 |
| exonElongation | 38 | | 0.395 | 0.342 | 0.263 |
| annotatedIntron_increasedUsage | 35 | | 0.171 | 0.771 | 0.057 |
| exonTruncation | 6 | | 0.833 | 0.167 | 0 |

**Figure S5:** Comparison of sequencing and event characteristics for FRASER2 events in ClinGenHI gene that did (n=78) and did not (n=70) pass manual verification

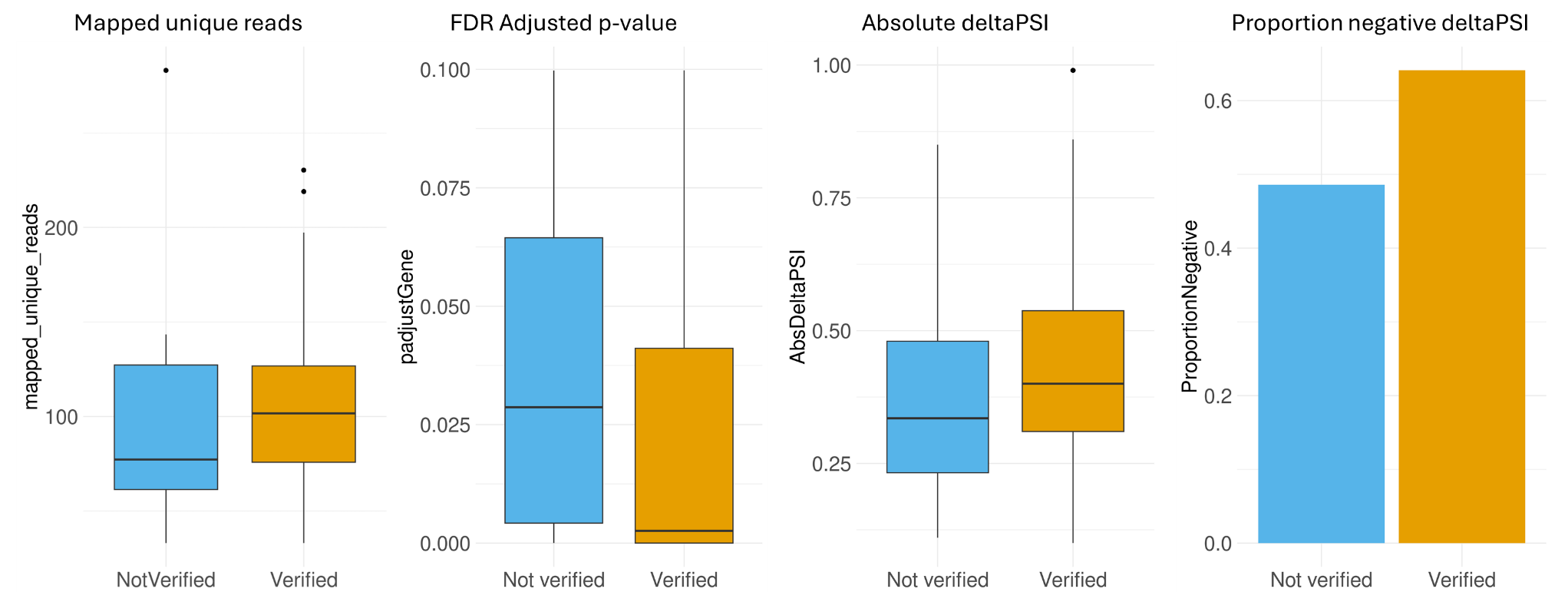

**Table S6:** Extended information on Case Study variants

| Case study no. | Gene | FRASER2 deltaPSI | FRASER2 p-adj | F2 raw event p | OUTRIDER fold change | OUTRIDER p-adj | OUTRIDER raw p | SpliceAI | gnomAD Allele Frequency |
| --- | --- | --- | --- | --- | --- | --- | --- | --- | --- |
| 1 | CTNNB1 | -0.32 | 0.000038242 | 1.08E-10 | 0.88 | 1 | 0.00037068 | NA | NA |
| 2 | PHIP | -0.48 | 0.0093043 | 1.4803E-09 | 1.09 | 1 | 0.01960324 | DL (0.98), AL (0.93) | 0 |
| 3 | WDR26 | 0.18 | 0.020018 | 1.1839E-08 | 0.91 | 1 | 0.16734877 | DG (0.83), DL (0.50) | 0 |
| 4 | KMT2D | -0.37 | 1.2188E-09 | 1.38E-16 | 1.39 | 2.12E-15 | 1.19E-20 | AL (0.27) | 0 |
| 5 | SPAST | -0.3 | 0.19427 | 3.94E-07 | 0.75 | 0.00026275 | 1.46E-09 | DG (0.66), AG (0.62) | 0 |
| 6 | RPL5 | -0.03 | 1 | 0.010507 | 0.56 | 2.82E-12 | 6.36E-17 | NA | NA |
| 7 | APC | -0.4 | 0.014499 | 2.46E-08 | 1.58 | 6.91E-25 | 3.91E-30 | NA | NA |
| 8 | FBXO11 | -0.45 | 0.1788 | 9.79E-08 | 1.32 | 3.21E-13 | 1.78E-18 | DL (0.95), AL (0.78) | 0 |
| 9 | INPPL1 | -0.61 | 2.8502E-06 | 6.09E-13 | 0.72 | 0.0003149 | 3.50E-09 | DG (0.52), DL (0.21) | 4.352E-06 |
| 10 | DYSF | -0.92 | 1.30E-49 | 1.16E-56 | 0.7 | 0.90397613 | 5.98E-05 | DG (0.42), DL (0.21) | 6.20E-07 |
| 11 | DNMT3A | -0.43 | 0.010778 | 7.8285E-09 | 1.09 | 1 | 0.05810143 | DG (0.89), AG (0.76), AL (0.26) | 9.379E-06 |

**Figure S6:**  SpliceAI and Pangolin predictions for splice disrupting variants

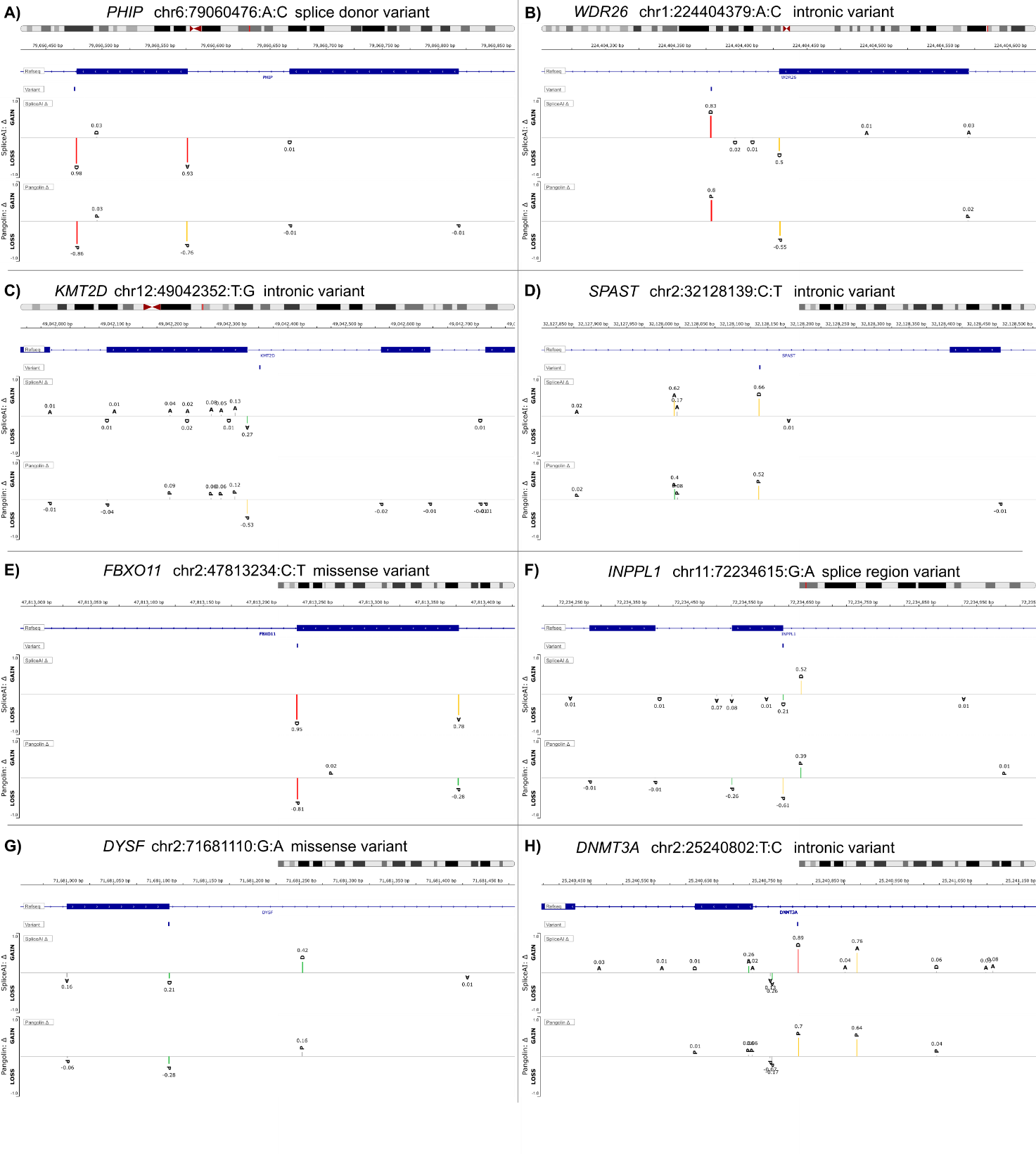

**Figure S7:** Example of pseudoexon creation in *SPAST*. A) bedGraph plots generated with SpliceAI-visual indicates that the NM_014946.4:c.1174-269C>T variant is likely to result in a 120bp pseudoexon (AG=0.62, -121; DG=0.66, -2). An interactive UCSC session is available at https://genome.ucsc.edu/s/AlistairP/SPAST_pseudoexon. The red rectangle highlights that the inserted sequence r.1173_1174ins120 contains two in-frame stop codons and so the predicted effect is a premature truncation, NP_055761.2:p.(A392Tfs*14). B) IGV derived Sashimi plot showing RNAseq data from two individuals heterozygous for NM_014946.4:c.1174-269C>T (P1-2) and 3 controls (C1-3). Confirms that the variant results in pseudoexon inclusion, consistent with the *in silico* prediction. P2 is the individual for whom OUTRIDER also gave a significant result (FC=0.75).

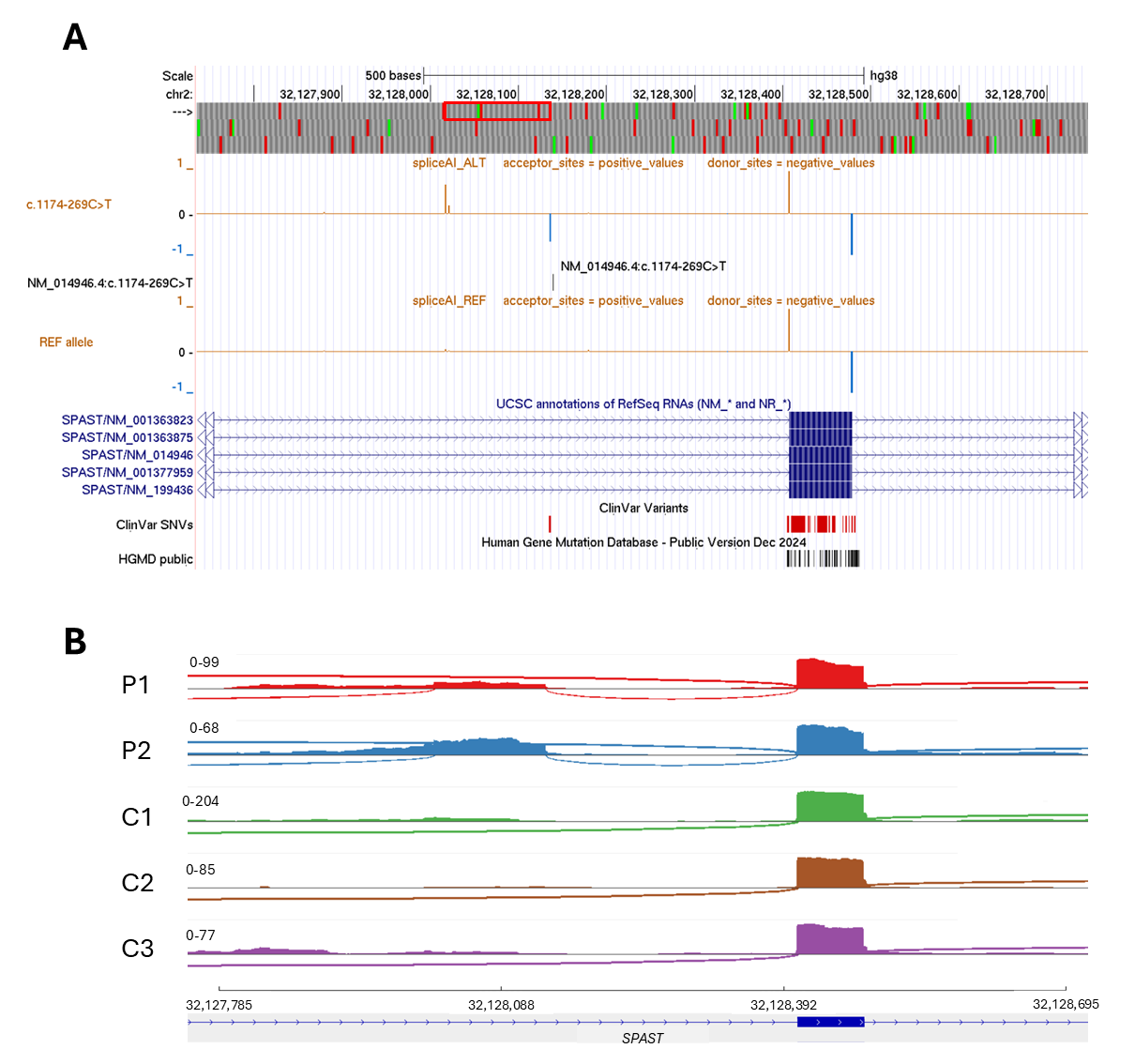

**Figure S8:** Read alignments showing *de novo* 5’-UTR deletion in *ANKRD11*. The top track (proband) shows reduced coverage and split read-pairs (highlighted in red) consistent with the presence of a 5.6kb deletion of *ANKRD11* exon 1. The same pattern is not seen in the father (middle) or mother (bottom) and so the variant has likely arisen *de novo*. Genomic coordinates are based on GRCh37 and shown in IGV using the “View as Pairs” option. The Canvas and Manta calls are plotted in light/dark blue, with the MantaDEL representing a more precise estimate of the true breakpoints.

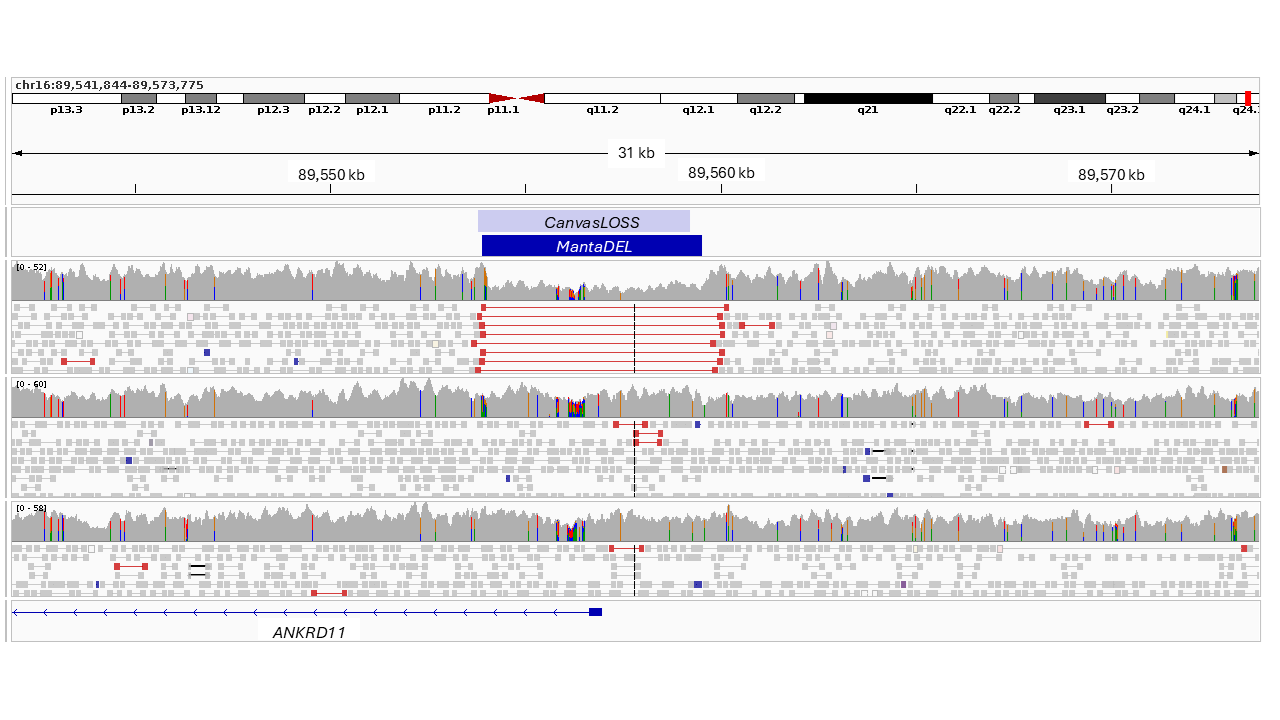

**Figure S9:** OUTRIDER outlier identified caused by deletion of first four exons of the *RPL5* gene. A) IGV screenshot showing RNA sequencing reads (above) and DNA sequencing reads (below). Red bars in DNA sequencing data indicate deleted region. B) Zoomed out view showing full extent of the proband’s deletion, spanning approx. 927kb. 7 additional protein coding genes fall within the deleted region. C) OUTRIDER adjusted p-values and fold change values for the genes within the deleted regions, plus the median TPM of the gene calculated across the cohort.

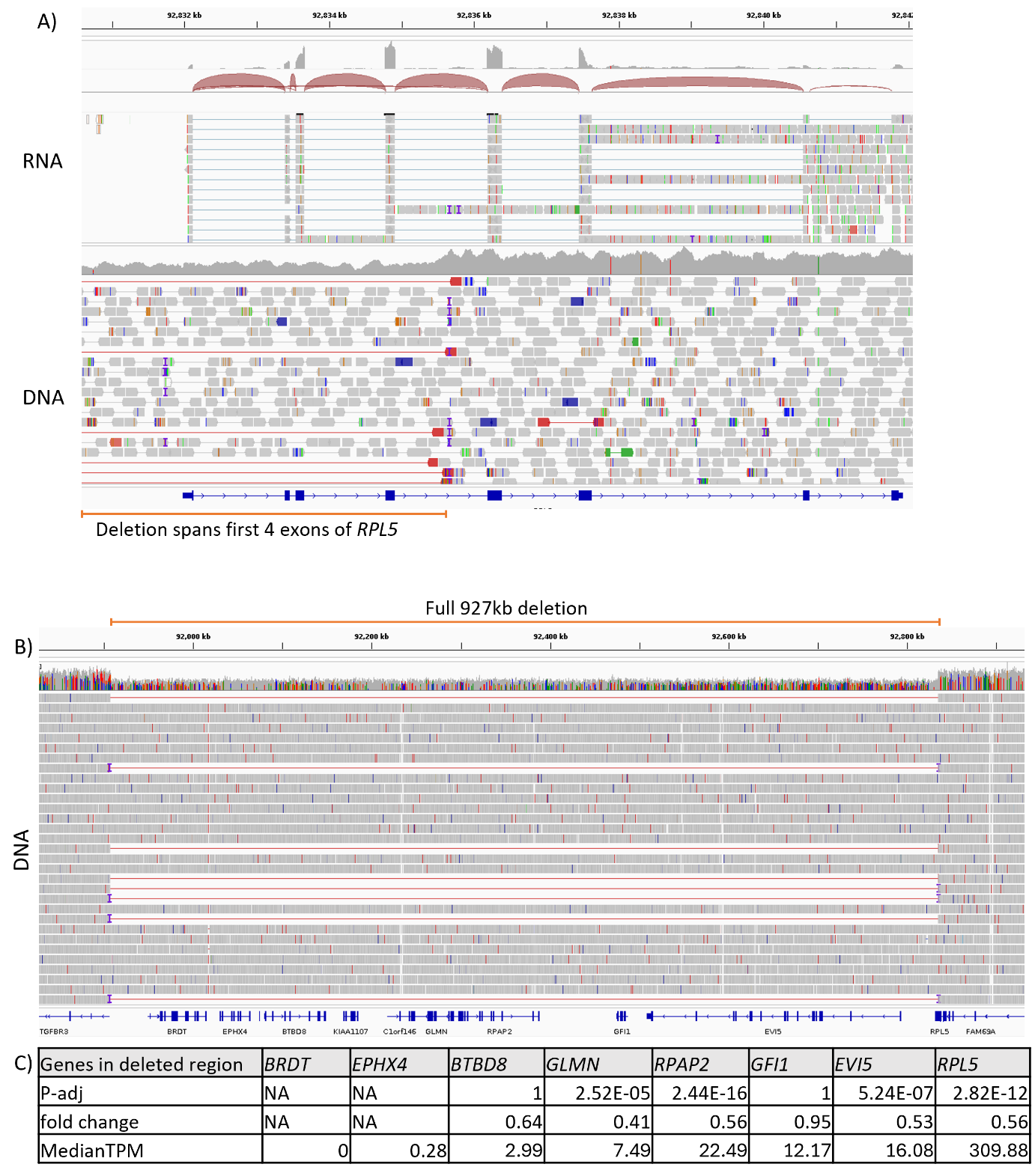

**Figure S10:** RNA and DNA sequencing reads of *APC* intron 10 in proband with multiple bowel polyps. Split reads indicate potential breakpoint, and evidence of PolyA insertion suggestive of retrotransposition event.

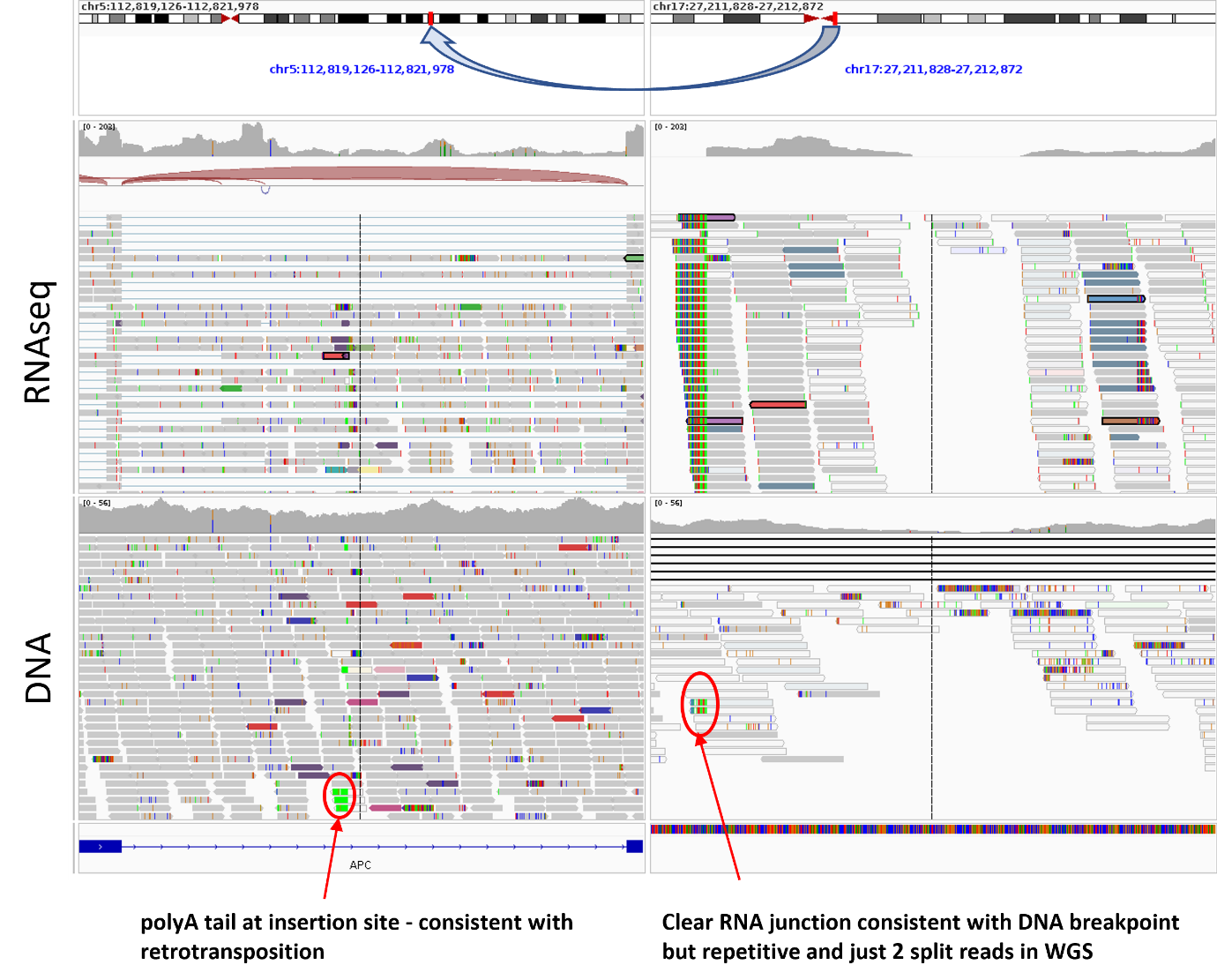

**Figure S11:** Nanopore long-read sequencing data for the *APC* locus confirms presence of SVA element that disrupts normal splicing. A) Long-read genome sequencing data showing presence of a 1.9kb insertion into intron 10 of the *APC* gene. Reads are separated into read-groups based on a common synonymous variant NM_000038.6:c.1635G>A (p.Ala545=). Due to the difficulties of correctly sizing homopolymer regions, the insert size is estimated at 1905-1932bp and matches closely the consensus SVA element. B) Long-read RNAseq data shows aberrant splicing is exclusive to the haplotype with an A at the polymorphic site in exon 14 (p.Ala545=), which is the haplotype with the SVA integration.

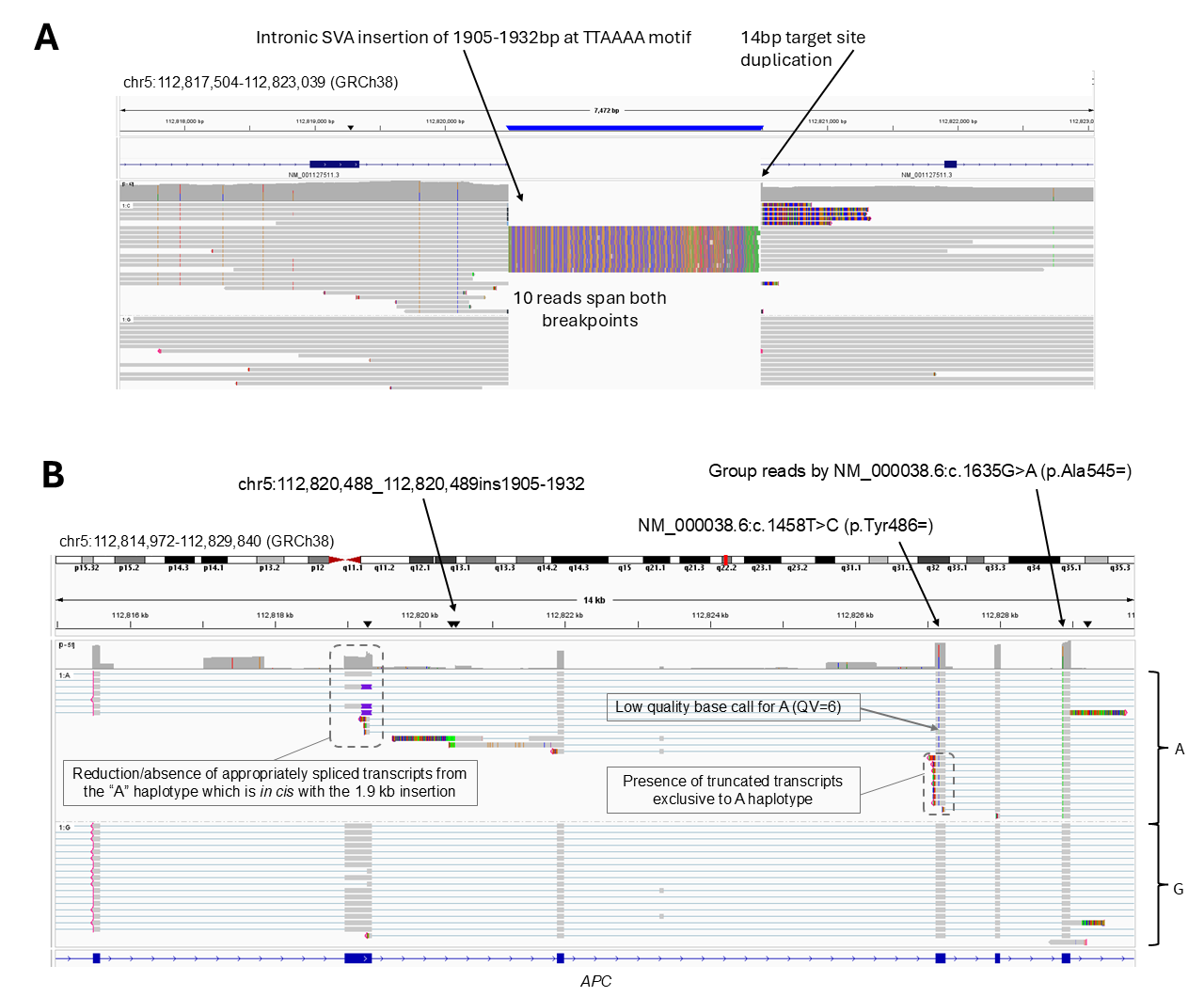

**Figure S12:** Sanger sequencing data confirming the breakpoints at each end of the SVA insertion. The insertion site lies in intron 10 of the *APC* gene (ENST00000257430.9/NM_000038.5), close to a TTTT/AA motif, which is known to be the preferred cut site for the L1 endonuclease.

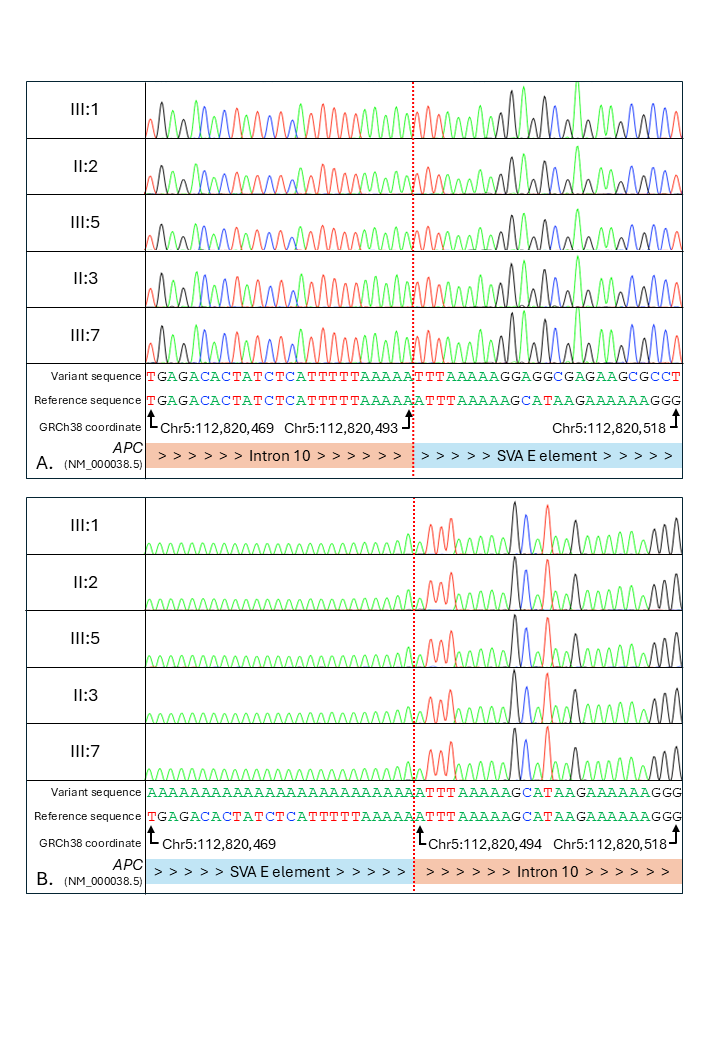

**Figure S13:** Cascade testing results for the SVA insertion in *APC*. A) Pedigree showing affection status for multiple bowel polyps (shaded symbols). Across 3 generations, there were 8 affected individuals of whom DNA was available for 5. The individuals highlighted in red were shown to harbour the SVA insertion. B) UV illuminated agarose gel picture showing the presence of a lower 291bp amplicon indicating the presence of the insertion. The lower band was not detected in a normal control DNA sample.

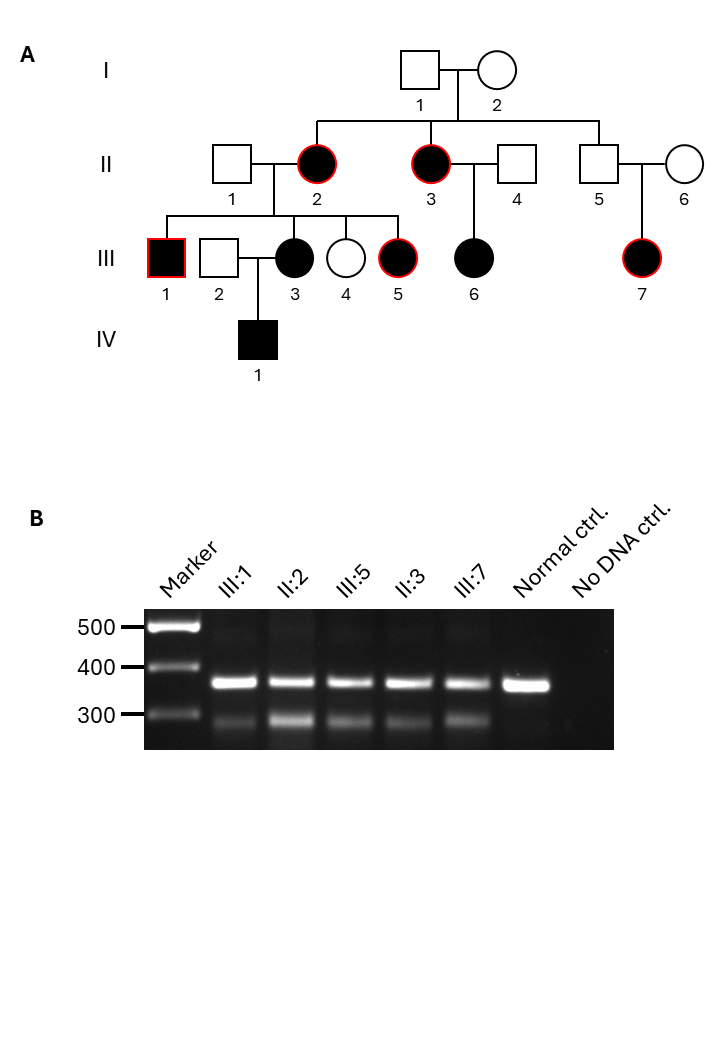
